# Predictive and Seasonal Dynamics of the Human Wastewater Virome

**DOI:** 10.64898/2026.03.19.26348845

**Authors:** Z. Vahdat, S.L. Grimm, T. Gandhi, M. Tisza, S. Cregeen-Javornik, S. Bel Rhali, J. Clark, H. Prakash, J. Petrosino, T. Ayvaz, M. Ross, J. Deegan, C. Bauer, E. Boerwinkle, C. Coarfa, A. Maresso

## Abstract

Wastewater-based epidemiology provides a scalable, noninvasive framework for population-level infectious disease monitoring, but traditional assays limit detection breadth and genomic insight. To address these constraints, we conducted targeted hybrid capture virome sequencing across 15 Texas cities over three years, from 2023 to 2025, generating ∼3 billion viral reads and identifying more than 900 strains across 374 species. Comprehensive temporal and spatial analysis revealed that the wastewater virome exhibits strong, predictable seasonal patterns, which grouped into three dominant seasonal clusters encompassing human, animal, and plant pathogens. Correlation network analysis revealed numerous positive co-occurrence patterns, including seasonal viral pairings, suggesting that the virome functions as a structured and interconnected ecological system. Leveraging this structure, we developed machine learning models using site-specific historical data to forecast individual viral species one month in advance. Of the 159 species modeled, approximately half achieved prediction performance of Pearson’s Correlation Coefficient R² ≥ 0.50, and many exceeded R² ≥ 0.75. Classification models accurately inferred the month and season of sample collection (AUROC > 0.85 and > 0.95, respectively). Predictive features frequently included other viruses and temporal indicators, highlighting networked, seasonal virome dynamics. Sentinel pathogens (e.g., Norovirus, SARS-CoV-2) could be forecast accurately even with limited historical data. Together, these findings demonstrate that the wastewater virome is highly seasonal, interconnected, and forecastable, providing a foundation for proactive, metagenomics-based monitoring and early outbreak detection.

## INTRODUCTION

Wastewater-based epidemiology (WBE) is a comprehensive, non-invasive, clinically independent mechanism for monitoring infectious diseases at the community level (Mao, Zhang et al. 2020, Kilaru, Hill et al. 2023, Deák, Lupu et al. 2026). The COVID-19 pandemic accelerated global adoption of WBE (Bivins, North et al. 2020, Stadler, Ensor et al. 2020, Amman, Markt et al. 2022, Bonanno Ferraro, Veneri et al. 2022, Prado, Rey-Benito et al. 2023). By analyzing viral genetic material shed into sewage systems, WBE provides near real-time insights into pathogen prevalence and transmission dynamics (Hata, Hara-Yamamura et al. 2021, Ahmed, Bivins et al. 2022, Boehm, Hughes et al. 2023), facilitates clinical monitoring, enables early outbreak detection, and gives public health stakeholders actionable information for health-promoting interventions (Mao, Zhang et al. 2020, Orive, Lertxundi et al. 2020). More recent WBE has expanded to other viruses (e.g. flu, RSV) and even to other pathogens (e.g. bacteria) (Bonanno Ferraro, Veneri et al. 2022, Boehm, Hughes et al. 2023, Prado, Rey-Benito et al. 2023, Smith, Maqsood et al. 2024, Justen, Rushford et al. 2026) but is primarily limited in total target selection due to constraints in multiplexing multiple targets (Martínez-Puchol, Rusiñol et al. 2020, Kilaru, Hill et al. 2023) targets. This reduces the total amount of data that reports on the “virome”, or by extension the “pathome” (all pathogens), present in a single wastewater sample (Nieuwenhuijse, Oude Munnink et al. 2020, Tisza, Javornik Cregeen et al. 2023). Such constraints also do not provide genomic context, which is important to monitor the emergence of new pathogens, their evolution towards greater transmission or virulence, and specificity in sub-classifying into lineages and strains (Karthikeyan, Levy et al. 2022, Schumann, de Castro Cuadrat et al. 2022, Yousif, Rachida et al. 2023) that allow for evolutionary tracking (Tisza, Javornik Cregeen et al. 2023, Wardi, Belmouden et al. 2024). Fully leveraging the potential of machine learning to detect emerging threats requires substantially larger and more complex training datasets (Nunes, Thommes et al. 2024).

To address these limitations, our group has used wastewater sequencing and metagenomics to infer pathogen presence, tracking, and evolution, a designation we term wastewater genomic epidemiology (or WGE) (Clark, Terwilliger et al. 2023). WGE allows for the same advantages offered by traditional WBE (e.g. the detection of pathogens and the reporting of their abundance levels), but also adds a comprehensive assessment of the entire pathogen matrix, thereby generating large-scale metagenomics information for deep analysis (Tisza, Javornik Cregeen et al. 2023). These approaches have allowed for the detection of “known unknowns” (such as measles outbreaks and avian flu emergence), as well as revealing novel spatial temporal relationships across the human virome (Tisza, Hanson et al. 2024, Javornik Cregeen, Tisza et al. 2025). Recent advances in machine learning (ML) and artificial intelligence (AI) have been applied to WBE for the purposes of performing a predictive analytics framework. This includes integrating heterogeneous data streams (e.g. viral concentrations, population demographics to forecast infection trends, coupling sewer network models to improve Covid detection (Zehnder, Béen et al. 2023), or use of time-series (Long Short-Term Memory – LSTM) as a regression approach to predict COVID case trajectories (Hu, Han et al. 2024). Such approaches have been also applied to forecast other pathogens such as influenza, RSV, mpox, and other pathogens.

Recognizing the ML-based approaches will aid in driving predictive models that may have real-world value to protecting populations from viral outbreaks, we report here the application of ML and correlative analysis to three years of continuous sequencing of wastewater from 15 different cities covering billions of different viral species reads. Our results demonstrate the human virome largely is self-predictive, demonstrating a total virome seasonality, while showing that multiple unexpected factors allow for viral forecasting, thereby offering novel parameters by which one may predict future virome and disease activity.

## RESULTS

### The Wastewater Virome

Since May of 2022, the Texas Epidemic Public Health Institute’s wastewater monitoring program (TexWEB) has been conducting agnostic hybrid-capture sequencing of all known human and animal viruses in cities across the state of Texas (Figure 1A) (Clark, Terwilliger et al. 2023, Tisza, Javornik Cregeen et al. 2023, Tisza, Hanson et al. 2024, Javornik Cregeen, Tisza et al. 2025). TexWEB includes 15 cities with a total population of ∼7 million people, 43 distinct catchment sites, and nearly 3,000 total samples sequenced and analyzed (Figure 1B). The hybrid capture enrichment panel utilizes over 1,000,000 oligonucleotide probes that match genomes from over 3,000 human, animal, and plant viruses. Through a period of 3 years of weekly to biweekly analysis, we have detected over 900 uniquely annotated viral strains belonging to 374 species across more than 100 genera (Figure 1C, D). Since its inception, the program has generated nearly 100 billion sequencing reads, of which 2.6 billion were unambiguously assigned to a viral taxonomy (Figure 1E). Seeking a deeper understanding of the data, the abundance of viral reads was analyzed in space and time using a combination of tools adapted from various fields. These include transcriptional gene responses (RPKMF), data hierarchical and structure clustering (Leiden clustering, BBKNN batch correction), association studies (Wilcoxon, Spearman, etc.) and machine learning (Random Forest) for prediction. The results of this analysis are presented herein.

**Figure 1.**
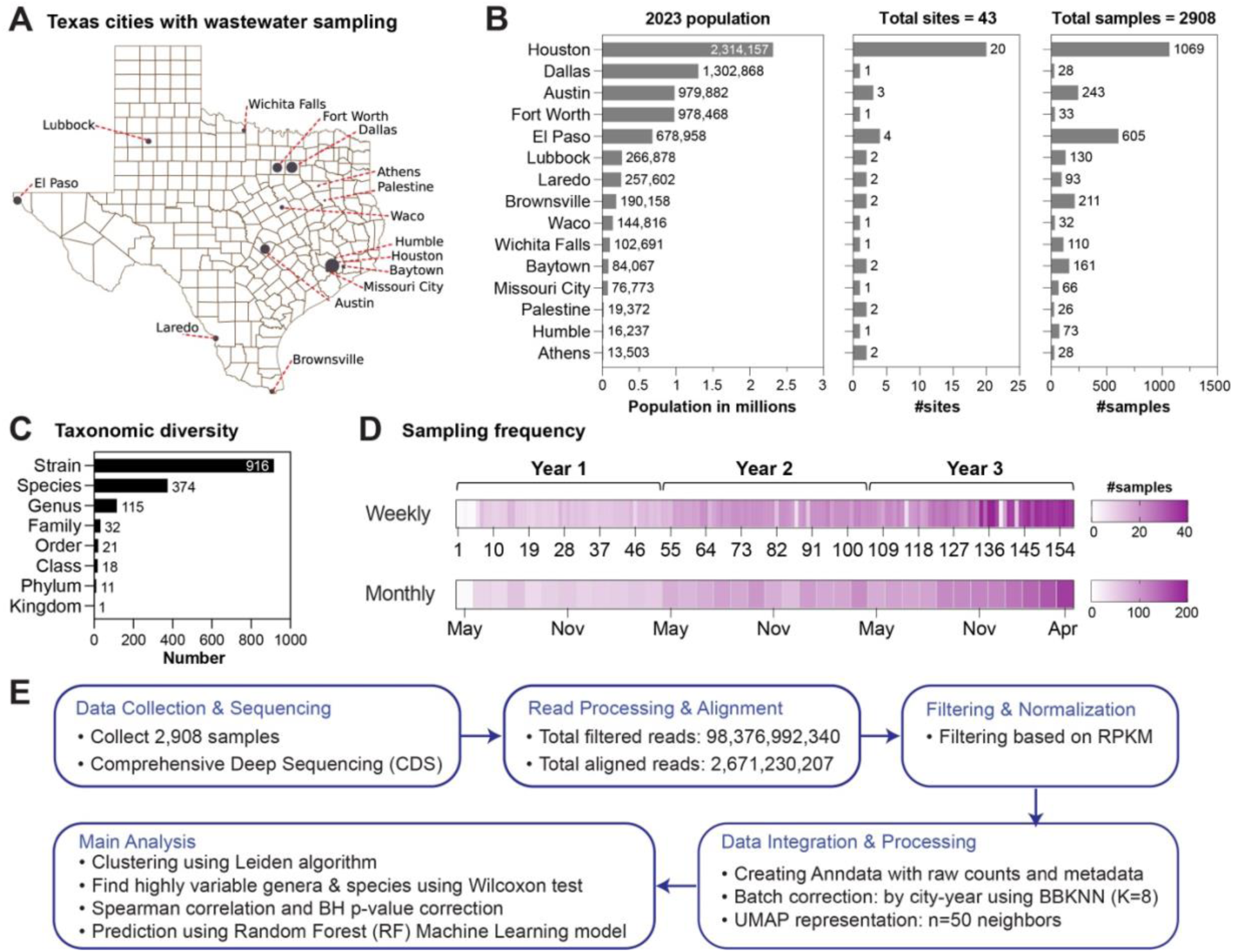
Overview of data generation and analytical approach. **A.** TexWEB conducted capture sequencing of the human and non-human virome in 15 cities across Texas with a total population of over 7 million. **B.** Detailed breakdown of the population in 2023, number of collection sites, and total samples collected in each city. **C.** Diversity of data collected reported at multiple taxonomic levels. **D.** The sampling frequency at weekly and monthly levels across the three years of the study presented. **E.** Overview of the analytical workflow, including data collection and sequencing, read processing, filtering and normalization, data integration. The main analysis includes clustering and cluster markers detection, determining highly correlated genera and species, and constructing predictors using machine learning for pathogen, sample collection month, and sample collection season.

### The Seasonality of the Virome

Utilizing three years of continuous and frequent sampling, we first examined the temporal nature of the virome in municipal human wastewater (WW). Plotting the RPKMF (Reads Per Kilobase per Million Filtered reads) values of each virus as a uniform manifold approximation and projection (UMAP) revealed robust organization in space and time (Figure 2). For example, the virome clusters by location, with data from a given city more closely related in time to itself than other geographic locations (Supplementary Figure 1A). This effect is partially mitigated by batch correction (Figure 2A), as evaluated by the kBet batch effect metric (Supplementary Table 1). This finding is consistent with our earlier observations and those of others that viral evolution is first locally restricted (Tisza, Javornik Cregeen et al. 2023). Additionally, there was consistent organization of the data in time (Figure 2B,C). Although there were clear patterns by year and month, the most striking observation was a strong seasonal separation of the data (Figure 2D). These seasonal distributions were not caused by any unusual unevenness in the data since batch uncorrected raw data showed the same effect (Supplementary Figure 1A-E). A deeper examination of the structure within the data showed that patterns were driven by certain clusters. After batch correction by city and year and using the Leiden community detection algorithm (Supplementary Table 1), as shown in Figure 2E, the data can be divided into at least three clusters with the greatest separation of data observed for a Leiden clustering at 0.4 resolution (Supplementary Figure 1F). Of the 115 genera detected in the data set, 88 showed strong organization in these three clusters (Figure 2F). Examination of the clusters clearly indicates that differences were driven by temporal trends (Figure 2G). Cluster c0 shows a more even distribution of viral signals throughout the year, with a rise in signal in the Fall months, a decrease through the Winter, and a noticeable absence of dozens of genera during the Summer (Figure 2G, H, I).

**Figure 2.**
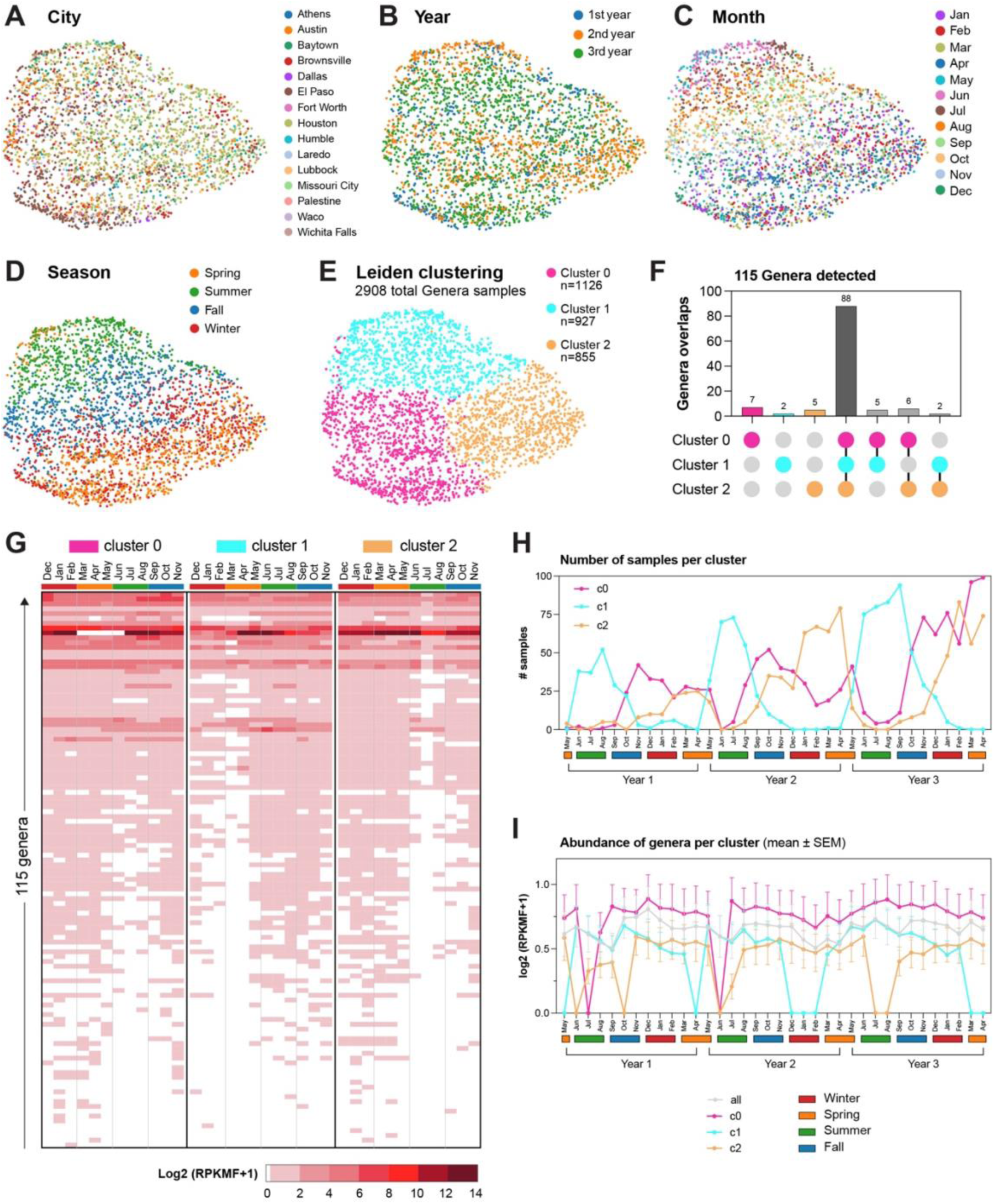
UMAP plots of viral genera showing the organization of samples by different **A.** cities, **B.** years, **C.** months and **D.** seasons after batch correction by city-year. **E.** UMAP plot showing the organization of samples divided by Leiden clustering at resolution of 0.4. **F.** A total of 115 unique genera was detected in the samples: 106 genera in cluster c0, 97 in cluster c1 and 101 in cluster c2, with 88 shared across all three clusters. The number of genera unique to each cluster and those shared between only 2 clusters are shown. **G.** Mean log-transformed RPKMF values for each of 115 genera across clusters, seasons and months. **H.** Number of samples assigned to each cluster across years, seasons and months. **I.** Mean log-transformed RPKMF of genera across years, seasons and months. The trend shows monthly averages with standard errors (SEM).

Rather strikingly, two additional clusters, Cluster c1 and Cluster c2, defined strong Summer and Winter signals, respectively. Cluster c1, for example, begins to rise in late Spring, peaks in Summer, and recedes in early Fall. In contrast, Cluster c2 begins to rise in late Fall, peaks in Winter, and recedes in early Spring (Figure 2G, H, I). The number of samples is lower in clusters c1 and c2 compared to cluster c0, which is more ubiquitous (Supplementary Figure 1G, Figure 3A). In both cases, more than 75% of the viral genera are absent in non-peak season for these clusters compared to cluster c0. Examination of the viral signal by month confirmed that cluster c1 showed reduced viral levels in December, January, February, March, and April, while the same was true for cluster c2 in June, July, and August (Figure 2H). The seasonal patterns of sample distribution per cluster and season aggregated over all wastewater collection sites (Figure 2H), is also observed at the level of individual cities (Supplementary Figure 2D). This remains evident even though cities with lower numbers of samples provide a coarser resolution of the seasonal patterns compared to Houston and El Paso. Further, in some cities with fewer samples, such as Athens, Dallas, Fort Worth, Palestine, and Waco, cluster c1 seems to shift upwards from a sample Summer peak to a Summer and Fall peak. As the program continues and more samples are collected, the distribution of samples per cluster in each city will be refined. This will allow for the evaluation of whether observed differences in temporal patterns are truly a city-specific phenomenon, or simply an artifact of differential sampling rates.

**Figure 3.**
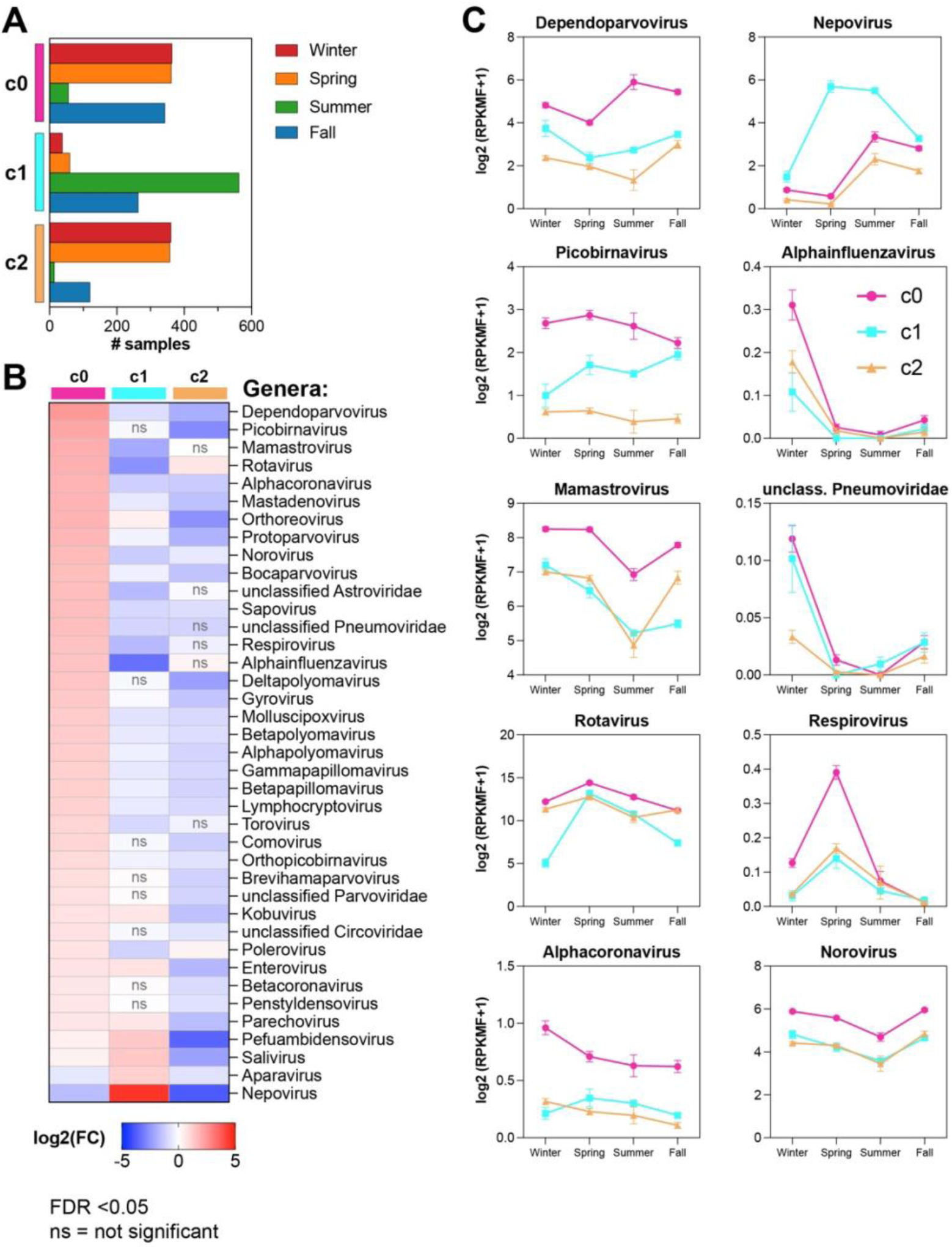
Genera markers of the three sample clusters and their seasonal distribution. **A.** Distribution of samples across seasons in each cluster. **B.** Selected top significant genera in each cluster, identified using the Wilcoxon test by comparing log-transformed RPKMF in one cluster against those in the other two clusters. **C.** Log-transformed RPKMF mean in each season for some of the top genera. Significant genera selected after filtering for FDR<0.05.

Based on the three clusters detected, we next assessed the species level resolution of the virome (Supplementary Figure 2A,B). Of the 374 identified species, 224 were shared across all 3 clusters (Supplementary Figure 2C). The aggregated species abundance (Supplementary Figure 2B) follows the temporal patterns of the aggregated genera (Figure 2I). Having established that samples in each cluster have a seasonal distribution (Figure 3A, Supplementary Table 3A), we determined the genera and species markers for each cluster along with the temporal distribution of these markers. Using a non-parametric Spearman rank test, we followed approaches commonly used in single cell analysis to identify and plot genera significantly increased in each cluster (Figure 3B). We then explored the distribution of these markers over the course of 4 seasons at the genera level (Figure 3B,C, Supplementary Figure 3, Supplementary Table 3B,C) and at the species level (Supplementary Figure 4, Supplementary Table 4). All identified genera and species were manually curated using publicly available databases and literature and classified as human or non-human pathogens. The complete list is provided in Supplementary Table 2.

Interestingly, an examination of the top ten viral genera driving the more ubiquitous signal in c0 indicates the majority of them infect humans (e.g. Norovirus), but can also infect other mammalian species (Mamastrovirus, Bocaparvovirus) or even other vertebrate species (Dependoparvorvirus, Rotavirus, Orthoreovirus, Protoparvovirus) (Figure 3B).

Strong seasonal genera included Nepoviruses (plant viruses which likely rise in Summer because of peak harvest season, highest in c1), the Alphainfluenzae (rise in Winter with flu season, highest in c0), Pneumonviridiae (RSV and hMPV – both causes of respiratory infections in people, high in Fall and Winter and in c0 and c1), and Respiroviruses (HPIV – causes respiratory infection, high in c0), as seen in Figure 3C. Additionally we have conducted a matched analysis of cluster markers and seasonal distribution at the species level (Supplementary Figure 4A-G). In general, these seasonal patterns held true on a species level and were independent of city or geography (Supplementary Figure 2C). A complete list of all genera and species including their levels over 3 years by season is shown in Figure 3, Supplementary Figures 3 and 4, and Supplementary Tables 3 and 4. Collectively, this data indicates the human and environmental virome (as comprehensively measured in unison via WW) displays a seasonal cycle.

**Figure 4.**
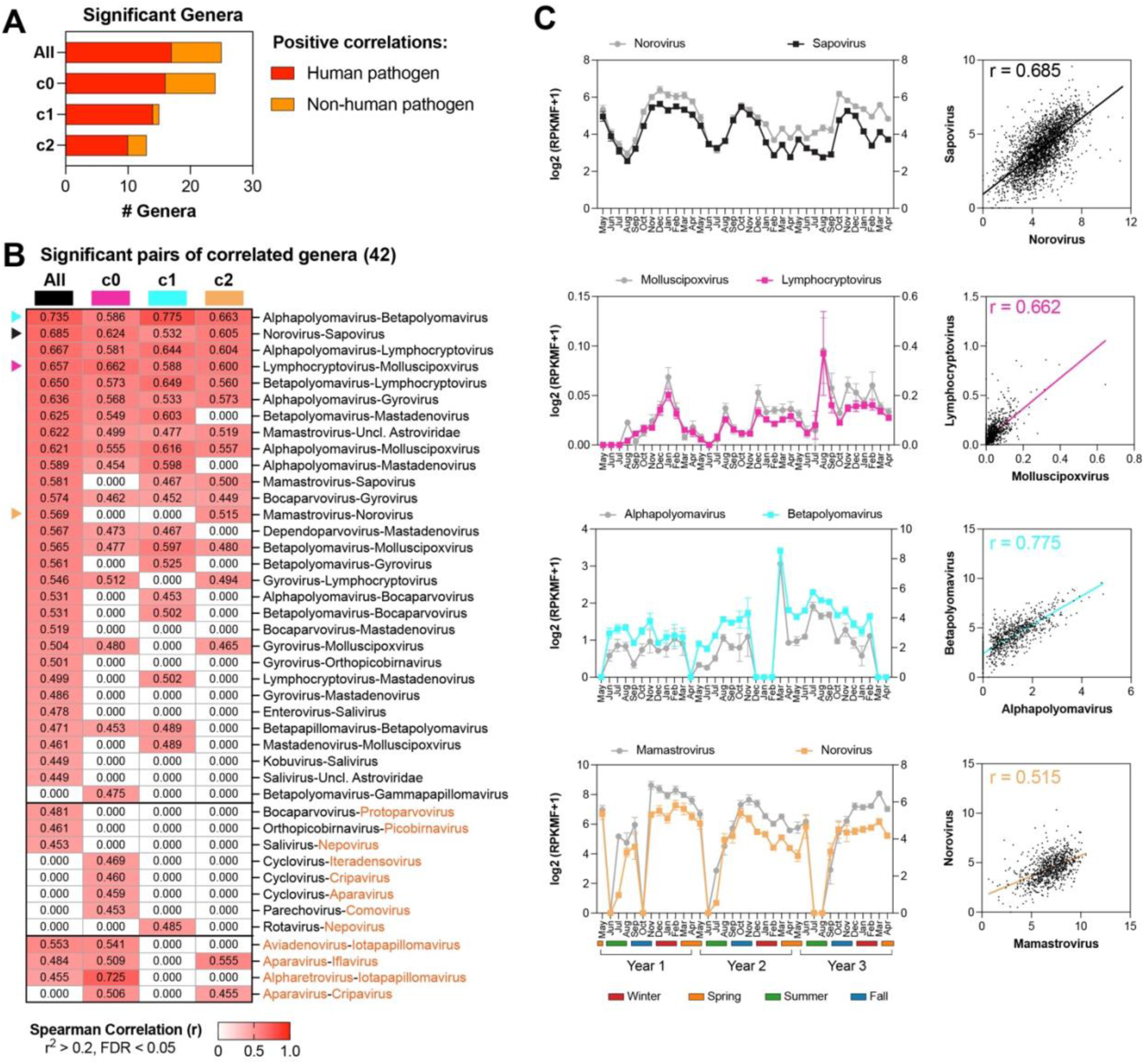
Virome correlation networks at genus level resolution. Spearman rank correlation was used to determined correlated genera, at FDR<0.05 and r^2^>0.2. **A.** Number of human and non-human genera that show significant correlations using the pooled samples or within each cluster. **B.** Heatmap showing the Spearman rank correlations for significant pathogen pairs across all pooled samples and in each individual cluster. Human pathogens are indicated with black font, non-human pathogens with orange font. **C.** Visualization of selected correlated pathogen pairs across 36 months and as scatterplots.

### Human, Animal, and Plant Pathogens Contribute to Virome Seasonality

Having established the seasonal and monthly patterns of detected clusters, including taxonomic markers for each (Supplementary Tables 3,4), we next explored pathogens exhibiting strong seasonal patterns. An exciting and quantitative finding is that pathogens capable of infecting different clades of the Tree of Life, spanning humans, animals, and plants, display seasonal patterns. We determined the high abundance genera and species within each season and associated them with the sample clusters where they predominate.

Among genera with *Spring peaks* specific to c0 are the human pathogens Picobirnavirus, Mamastrovirus, Respirovirus, unclassified Astroviridae, and Rotavirus (shared with c2) (Figure 3B,C, Supplementary Figure 3A). Non-human pathogens with Spring peaks in c0 are Penstyldensovirus and Polerovirus (shared with c2) (Figure 3B,C, Supplementary Figure 3B). Penstyldensovirus infects prawns and shrimp, whereas Polerovirus infects cereal, vegetable, and fruit plants. Observed temporal patterns of these pathogens might reflect dietary consumption of the respective food groups.

At species resolution, c0 pathogens with Spring peaks included Mamastrovirus 1, Human astrovirus, Human respirovirus 3, and Rotaviruses A and C (shared with c2) (Supplementary Figure 4A,B,E,F). The cluster c0 also contains viruses that are commonly associated with human clinical samples, such as human astrovirus, discovered in 1975 in children with gastroenteritis. Its continuous presence in the sewershed likely reflects its high prevalence in human populations. Non-human pathogens in c0 exhibiting Spring peaks included Porcine picobirnavirus, which is also high in Summer, and Decapod penstyldensovirus 1, which affects shrimp. A non-human Spring peak pathogen high in both c0 and c2 is the Cucurbit aphid-borne yellows virus, which infects cucurbits including melons, squash, and cucumbers, leading to yellowing of the leaves (Supplementary Figure 4A, B,G).

Examining the genera driving Summer peaks led to interesting findings. Of the 9 significant genera showing a Summer peak in cluster c1, 5 are viruses associated with humans and animals (Figure 3B,C, Supplementary Figure 3B, Supplementary Table 3B). This includes Saliviruses (SaV-A1 etc.), human Enteroviruses (EnvA/B/C), Kobuviruses (AichivirusA or AiV-A), Paraechovirus (HpeVA), and Orthoreoviruses. All these viruses cause human gastrointestinal or respiratory infections, some serious, with HpeVA causing meningitis in children. An interesting finding for Summer was the rise of Aparavirus, which can cause paralysis in bees (Supplementary Figure 3B), and Nepovirus which infects tomatoes (Figure 3C), likely reflecting increased pollination activities as plants bloom and gardens mature in the warmer months. Despite c0 having a lower presence in Summer, it contains genera that are most abundant in Summer. The human-related genera include Dependoparvovirus, Enterovirus, Gyrovirus, Betapolyomavirus, Mastadenovirus, and unclassified Parvoviridae. Interestingly, Salivirus, Kobuvirus, and Parechovirus are also highest in c0 Summer and are shared as markers with c1. One non-human genus associated with both c0 and c1 and most abundant in the Summer is Pefuambidensovirus, which infects arthropods. This signal may relate to contribution to the sewershed input by insects.

At the species level, human pathogens specific to c0 with Summer peaks include Adeno-associated dependoparvovirus A and B, Human mastadenovirus B, D, and F, Betapolyomavirus hominis, Betapolyomavirus secuhominis, and Mamastrovirus 8 and 9 (Supplementary Figure 4A,B,E,F, Supplementary Table 4A). Summer peak pathogens shared by c0 and c1 include SARS-CoV2, Salivirus A, Salivirus FHB, Enterovirus C, Kobuvirus sp., and Parechovirus A (Supplementary Figure 4E,F). Finally, one Summer peak human pathogen specific to c1 is Astrovirus VA3. The non-human realm is very diverse (Supplementary Figure 4B,G). Representatives including Chicken anemia virus, which infects chickens, and Parus major densovirus, which infects insects and crustaceans., are more abundant in c0 Summer. Tobacco ringspot virus and the Solenopsis invicta virus 1, which infects the imported red fire ant Solenopsis invicta are high in c1 Summer (Supplementary Figure 4C,G). Finally, common to both c0 and c1 is Blattodean pefuambidensovirus 1 which infects the German cockroach.

The genera with *Fall* peaks are mostly c0 and c1 markers (Figure 3B,C, Supplementary Figure 3). Human pathogens specific to c0 with high abundance in Fall are Norovirus, Betapolyomavirus, Mastadenovirus, Sapovirus, and unclassified Circoviridae, while the human pathogen Parechovirus is shared with c1. Non-human pathogens in c0 with peaks in the Fall include Brevihamaparvovirus which infects mosquitoes, Comovirus which infects legumes, such as beans, peas, and soybeans, and Protoparvovirus that can infect dogs and cats.

The genera with Winter peaks are predominantly c0 markers (Figure 3B,C, Supplementary Figure 3). They include well known Winter pathogens Alphainfluenzavirus, Alphacoronavirus, Betacoronavirus, and unclassified Pneumoviridae, but also contain a wide variety of other pathogens such as Mamastrovirus, Norovirus, Alphapolyomavirus, Deltapolyomavirus, Bocaparvovirus, Betapapillomavirus, Gammapapillomavirus, Lymphocryptovirus, Molluscipoxvirus, and Torovirus. In the non-human realm, Protoparvovirus has high prevalence in Winter and Fall and can infect pets including dogs and cats.

At the species level, we observed many enteric pathogens high in c0 Winter including Norwalk virus, Mamastrovirus 1, Rotavirus I, Astrovirus MLB1, Astrovirus MLB2, Astrovirus MLB3, Human mastadenovirus B, Orthopicobirnavirus hominis, and Human mastadenovirus C and E (Supplementary Figure 4A,B,D,E,F). We also detected other common Winter pathogens high in c0: Alphacoronavirus 1, Influenza A virus, Rhinovirus C, Respiratory syncytial virus, Betacoronavirus 1, and Human coronavirus 229E (also high in c2). In the non-human pathogen realm, the highly abundant Winter virome included Feline rotavirus, Canine astrovirus, Carnivore bocaparvovirus 1 and 2, Carnivore protoparvovirus 1, and Porcine torovirus, all highly abundant in c0 (Supplementary Figure 4A,B,G). A full species list by cluster and seasonal abundance is shown in Supplementary Table 4.

### Elucidating the wastewater virome correlation network

Our large and robust virome abundance dataset enabled the construction of the virome correlation network, resolved at both genus and species levels. To identify pathogens with similar temporal patterns, we performed Spearman correlation analysis for all distinct pathogen pairs, followed by Benjamini-Hochberg multiple testing correction. Highly correlated pathogen pairs at the genus and species level were selected based on an FDR lower than 0.05 and a squared correlation coefficient (r^2^) exceeding 0.2 (e.g. r>0.45). This stringent filtering was needed to eliminate spuriously correlated pathogen pairs, particularly at the species level. In addition to building correlation networks across all samples, we also computed them within each sample cluster reported above and determined correlated pathogen pairs detected only in specific clusters. We further characterized our network based on the human or non-human pathogen determination(e.g. how many correlated pathogen pairs are in the human/human, non-human/non-human, or even human/non-human realms).

We first constructed genus-level virome correlation networks. With all data combined, we detected only positive genus-genus interactions. The interactions were comprised of 25 unique genera including 17 genera that can infect humans and 8 genera of viruses that don’t infect humans (Figure 4A). Across all combined samples, we observed 35 positively correlated genus pairs; three between non-human genera, three between human and non-human genera, and 29 between human pathogens (Figure 4B). A hypothesized benefit from determining three sample clusters is it may enable discovery of virome correlations that would otherwise be missed by pooling all the samples. We next computed the virome correlation network within each of the three clusters c0, c1, and c2. Figure 4A shows the number of unique genera involved in positive correlations within each cluster; 24 in cluster c0 (16 human and 8 non-human), 15 in cluster c1 (14 human and 1 non-human) and 13 in cluster c2 (10 human and 3 non-human).

Clustering revealed seven additional genus-level correlated pairs, resulting in a total of 42. In cluster c0, we identified one correlated pair between human genera (Betapapillomavirus and Gammapapillomavirus) and four correlated pairs between human and non-human genera–Cyclovirus with Iteradensovirus, Cripavirus, Aparavirus, and Parechovirus with Comovirus (Figure 4B). We also detected one additional correlated human-nonhuman pathogen pair in cluster c1 (Rotavirus with Nepovirus), and one correlated non-human pair in both clusters c0 and c2 (Aparavirus and Cripavirus). Across all samples, c0, c1, and c2, the majority of the virome correlations involved human/human pathogens; all samples, c0, and c2 show some non-human/non-human pathogen correlations (Supplementary Figure 5). Strikingly, there are very few human/non-human pathogen correlations at genus level (Supplementary Figure 5), with c1 having almost exclusively human/human pathogen correlations.

In addition to identifying correlations not detected in the overall dataset, clustering also revealed stronger correlations between certain pathogen pairs within specific clusters. In Figure 4C, we depicted four genus pairs with the highest correlations across the full combined data set (Norovirus/Sapovirus) or within clusters c0 (Molluscipoxvirus and Lymphocryptovirus), c1 (Alphapolyomavirus and Betapolyomavirus) and c2 (Mamastrovirus and Norovirus), respectively. We depicted the correlations using a scatterplot across all observations displayed their temporal patterns by plotting monthly average and standard error of means (SEM) across the three years of project collections. Despite the different ranges of each pathogen, the temporal patterns show striking similarities. Such similarities can be explained by co-infections, especially in immunocompromised individuals, but also by having matched seasonality and similar risk factors. Reported examples include Norovirus and Sapovirus (Umair, Rehman et al. 2023) and Norovirus and Astroviruses such as Mamastrovirus (Fernandez-Cassi, Martínez-Puchol et al. 2020).

Using a similar approach as described above, we computed the virome correlation networks at species resolution. Across all the pooled samples, we identified 50 species with positive correlations (27 human and 23 non-human) and two species with negative correlations (both human pathogens) (Supplementary Figure 6A). In total, 81 virome species pairs showed positive correlations – 50 human/human pathogens, 15 human/non-human pathogens, and 16 non-human/non-human species (Supplementary Figure 6B).

Using the sample clustering enabled the detection of additional correlated pairs that were not observed when pooling all samples. In cluster c0, 55 distinct species with positive correlations were identified, including 30 human and 25 non-human pathogens (Supplementary Figure 6A). In cluster c1, 38 distinct species with positive correlations were found, 26 human and 12 non-human pathogens. In cluster c2, 31 species with positive correlations were found, including 18 human pathogens.

In cluster c0, we identified eight new human/human pathogen pairs, three new human/non-human pairs and eleven non-human/non-human pairs, unique to this cluster (Supplementary Figure 6B). In cluster c1, we observed four human/human pairs and one human/non-human pair (Rotavirus A and Tobacco ringspot virus) unique to this cluster (Supplementary Figure 6B). In cluster c2, we identified one human with human (Mamastrovirus 9 and Sapovirus sp.), and two non-human pairs (Porcine torovirus and Bovine torovirus) and (Deformed wing virus with Varroa destructor virus 1), all unique to this cluster, (Supplementary Figure 6B). Across clusters, we found one nonhuman-nonhuman pair (Dragonfly associated cyclovirus 2 and Spodoptera exigua iflavirus 1) shared between cluster c0 and c1. Two non-human pairs, Rhesus adenovirus 58 with Murine astrovirus, and Solenopsis invicta virus 1 with Spodoptera exigua iflavirus 1, were shared between clusters c0 and c2 (Supplementary Figure 6B).

Supplementary Figure 6C shows the patterns of five representative species pairs over time, along with scatter plots illustrating the correlation of each pair. The figure highlights the temporal patterns of species either across the full data or within clusters c0, c1 and c2. Unlike genera, virome networks at species resolution reveal that while the majority of correlations are among human/human pathogen pairs, there are robust non-human/non-human correlated pathogens pairs (all samples, c0, c1), but also robust human/non-human pathogen correlations (all samples, c0, c1, c2). The cluster c0 harbors the largest number of non-human to non-human pathogen correlations.

### Developing predictors based on the wastewater virome using machine learning

Since the start of the TexWEB program, the wastewater sampling frequency increased, while the number of sampling sites increased substantially (from two sites in May 2022 to 43 sites in April 2025 (Figure 1D, 5A)). A tantalizing opportunity presented by this corpus of data was to develop predictors of relative abundance for each individual virus species using machine learning. In this study, using the wastewater information collected at each site, we developed predictors of relative abundance for each individual virus species one month in advance at the same site. When building these predictors, we tackled an additional question: how many months of history at one site are needed for optimal predictive performance, considering a range of one to twelve months of history. For a virus v, and using n months of complete virome abundance history (n in the range of 1 to 12), we developed predictors P(v,n); for a given site S, using n months of history, we would apply the predictor P(v,n) and determine the relative abundance of v at the site S in the following month. We modeled the time at which virus levels were predicted as individual months, e.g. January through December, or seasons Winter through Fall, but also as a numeric value, 1 through 12 for months and 1 through 4 for seasons. Finally, given the strong seasonality observed in the data, we developed predictors to determine the month and the season a sample was collected in. The setup of these machine learning problems is indicated in Figure 5C.

**Figure 5.**
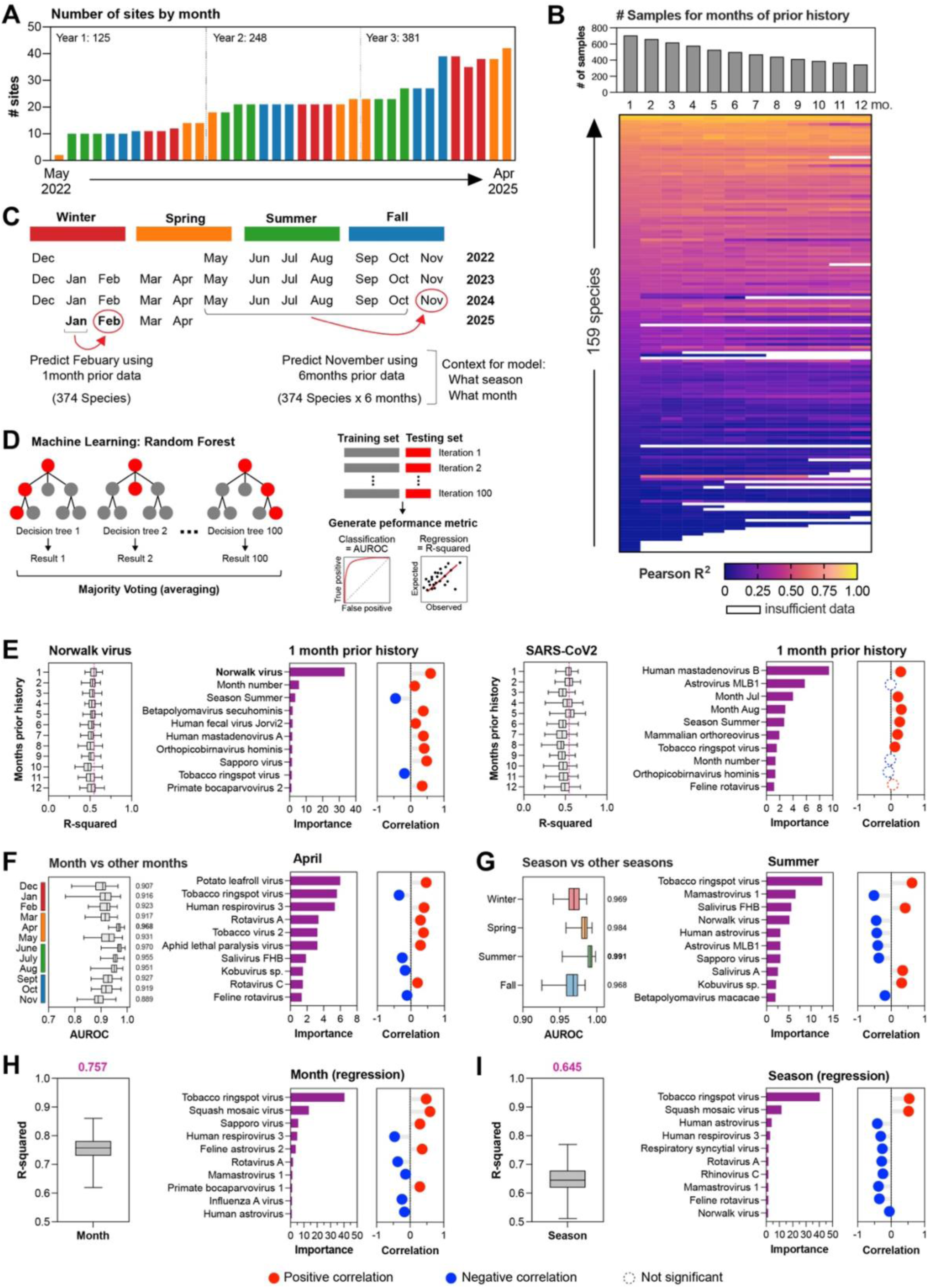
Developing machine learning predictors for pathogens, month, and season using the wastewater virome. **A.** Evolution of number of sites across the three years of the project. **B.** Summary of goodness-of-fit R^2^ predictors performance for virus levels one month in advance, using one to twelve months of history. **C.** Setup of predictors for virus levels one month in advance, for each site. **D.** Schematic of our Random Forest machine learning approach. **E.** Predictive performance R^2^ across the 100 cross-validation iterations using one to twelve months of history per site for the Norwalk and SARS-CoV2 viruses, and the top ten informative features for each together with their individual correlation with the predicted viruses. **F.** Prediction performance for each individual Month of sample collection, and the top informative features and direction of association with April. **G.** Prediction performance for each individual Season of sample collection, and the top informative features and direction of association with Summer. **H.** Prediction performance for Month as a numerical variable, the top ten informative features, and direction of association. **I.** Prediction performance for Season as a numerical variable, top ten informative features and direction of association.

Given the non-uniform number of samples collected over time (Figure 1D), as input for machine learning we used the averaged levels of viruses in each sample per month at each site. We utilized RandomForest (RF) classifiers for predicting individual months or seasons, and regressors for predicting month or season as numbers, or virus abundance one month in the future as a numerical continuous variable. RandomForest machine learning first develops a randomized collection of decision trees, then applies a majority voting scheme (Figure 5D). To enable scaling for our sizeable project, both in terms of samples and features (e.g. over 2900 samples and 374 species) we used RandomForest as implemented in the PyCaret and sklearn libraries. To determine robustly the performance of predictors built using our data corpus, we employed cross-validation as follows: in each iteration, input data was split into 70% training and 30% testing, a predictor was developed using RF, then it was applied to the testing data. For regression, the performance metric goodness-of-fit R^2^ (using Pearson Correlation) was computed in the test data, whereas for classification we utilized the area under receiver operating characteristic curve (AUROC). Finally, performance was aggregated over 100 cross-validation iterations, computing the median R^2^ or median AUROC (Figure 5D). To make our models transparent and interpretable, for each regression and classification model, the important features are reported in Supplementary Tables 5-7. We examined the top ten important features that appeared in at least 70% of iterations and computed the Spearman correlation between each important feature and the modeled variable, using one month history.

For predicting virus abundance for each species one month in advance, we summarize our RF performance results using 1 to 12 months of history in Figure 5B, indicating the median R^2^ across 100 cross-validation iterations. As the required site history increased from 1 to 12 months, the number of samples per site with sufficient history decreased (Figure 5B, top). For example, to predict Norwalk virus abundance levels for month November using six months of prior data (May to October), data were available from only 2 sites in 2022, 18 sites in 2023 and, 23 sites in 2024 (total 43 sites). As such, with only one month of history we used over 600 samples, but with twelve months of history we could only use 400 observations. Even though we quantified confidently 374 species across all samples, for each machine learning regression problem we required that a species has at least 20 non-zero values, leading to a total of 159 species for which we could develop predictors. In the heatmap from Figure 5B, we indicate with blank cells species/months of history pairs with insufficient data.

The heatmap in Figure B shows the ML regression results predicting virus abundance one month in the future. A striking pattern emerged, showing that for roughly half of the 159 species prediction achieved strong performance with R^2^ over 50%, whereas for the bottom half only a moderate to poor prediction is achieved. Among the top 30 species in the heatmap, some species show prediction correlations that remain constant despite increasing the length of training history. These species include Betapolyomavirus macacae, Tobacco ringspot virus, Rotavirus I, Rotavirus A, Alphacoronavirus 1, Betapolyomavirus secuhominis, Sapporo virus, Rhesus adenovirus 66, Mamastrovirus 1, Mamastrovirus 9 and, Sapovirus sp. In contrast, predictive performance R^2^ for Rhesus adenovirus 58, Blattodean blattambidensovirus 1, Salivirus FHB, Human fecal virus Jorvi2 slightly increased when the history length is extended to two months or more. For some species, R^2^ increased slightly with training histories of 2-8 months and then decreased. These include Human mastadenovirus B, Human mastadenovirus G, Astrovirus VA3, Burkina Faso astrovirus, Astrovirus MLB3, Canine astrovirus, Squash mosaic virus, Carnivore protoparvovirus 1, SARS-CoV2, Astrovirus MLB1. We expect that this decline is due to the reduction of training samples with longer histories available, rather than a limitation in the prediction power of the model. A similar trend is observed for Feline rotavirus, Human respirovirus 3, Human astrovirus and Norwalk virus where R^2^ slightly decreased by increasing history beyond one month.

In the middle and lower parts of the heatmap in Figure 5B, with modest to moderate predictive performance, we observed many species whose R^2^ values increased as the training input history extended from 1 month to 2-12 months. These species include Orthopteran scindoambidensovirus 1, Salivirus A, Canine morbillivirus, Potato leafroll virus, Influenza A virus, Human coronavirus 229E, Gammapapillomavirus sp., Dipteran protoambidensovirus 1, Callinectes sapidus reovirus 1, Human coronavirus HKU1, Callinectes sapidus reovirus, Cardiovirus D, Rotavirus B, Monkeypox virus, Murine mastadenovirus B, Cardiovirus B, Human betaherpesvirus 7, Human respirovirus 1, and Yongsan iflavirus 1.

Based on general interest and potential impact on community health, we selected 14 human pathogens among the 159 modeled species. The R^2^ values for Norwalk virus and SARS-CoV2 across 1-12 months of training input history are shown in Figure 5E. For both pathogens, the median R^2^ slightly decreased as the training history length increased, suggesting that 1-month of training history is sufficient to build the model; however, by achieving R^2^ of 50%, both viruses can be modeled with strong predictive performance. The most recurrent important features for the models with 1-month of training history are listed in Supplementary Table 5. When modeling Norwalk virus levels with 1-month history, we found that abundance of Norwalk virus itself was the most important feature, showing a strong positive correlation with its own abundance one month in advance. Interestingly, the month and season for which Norwalk virus abundance was predicted were the second and third most important features, respectively (Figure 5E). Seven of the remaining top ten features were human pathogens, all positively associated with Norwalk virus levels. These results are in agreement with the general understanding that Norovirus infections are present throughout the year. For SARS-CoV2, the top ten important features are shown in Figure 5E. Using a 1-month prior history for model training and testing, we found that 4 of top ten features were human pathogens; Mammalian orthoreovirus and Orthopicobirnavirus hominis are positively associated with SARS-CoV2 abundance one month in the future. The months of July and August, and the summer season were also identified as important temporal features contributing to SARS-CoV2 model construction, as potential inflexion points in community spreading. These results corroborate our understanding of seasonality of SARS-CoV2 in Texas.

The remaining 12 human pathogens of high public interest along with their R^2^ values across months, the top ten important features from the 1-month history models, and the direction of association between each feature and the modeled pathogens abundance level, are presented in Supplementary Figure 8. The full list of the most recurrent features is in Supplementary Table 5. For 6 of the 12 pathogens, Rotavirus A, Enterovirus B, Enterovirus D, Human mastadenovirus B, Primate erythroparovirus 1, Primate norovirus, the most important feature when using one month of history is the same virus, indicative again of the ubiquitous presence of these pathogens throughout the year. Among this group, prediction of Rotavirus, Enterovirus D, and Human mastadenovirus B achieves R^2^ at or above 50%. For the remaining six viruses, Human respirovirus 1, Human respirovirus 3, Influenza A, Respiratory syncytial virus, Monkeypox virus, Hepatovirus A, which have strong seasonality, the most important feature is not the virus itself, but interestingly other viruses, perhaps indicating seasonality. Of note, a recurrent top ten predictor among Norwalk virus, negatively associated with Enterovirus D, and positively associated with six of the 14 viruses of interest. Another striking observation is the presence of time indicators, particularly month number which was positively correlated with Human mastadenovirus B, Norwalk virus, and Respiratory syncytial virus, but negatively correlated with Influenza A, Rotavirus A, and SARS-CoV2. However, individual months or seasons associate with individual pathogens too, such as month of January for Influenza, September for Enterovirus D, and Spring for Human Respirovirus 3. Overall, our prediction of pathogen levels shows that it can be conducted robustly, but different classes of pathogen have highly distinct emergence patterns and co-occurring or mutually exclusive species based on the correlations reported for the informative top ten features.

Based on the strong seasonality observed in the data (Figure 2C,D,H,I) we surmised that given any sample predictors could be developed determining the month and season that sample was collected. First, RF classifiers were developed models for each month (e.g. January through December) and each season (Winter through Fall). The list of top important features for each season and month is in Supplementary Table 6. The median prediction AUROC was above 0.85 for months (Figure 5F) and above 0.95 for seasons (Figure 5G), indicating strong predictive power for determining whether a sample belongs to a specific month or season. The top ten important features for April and for the Summer season are shown in Figure 5F. The top ten features for all months and seasons are in Supplementary Figure 9.

For predicting the month of April, among top 10 features, Human Respirovirus 3, Rotavirus A, and Rotavirus C were positively associated human pathogens, while Salivirus FHB and Kobuvirus sp. were negatively associated human pathogens. Interestingly, some non-human pathogens are highly informative for April, specifically Potato leafroll virus, Tobacco virus 2 with higher abundance in April (e.g. positive correlation) and Tobacco ringspot virus and Feline Rotavirus, which have higher abundance in the months other than April. For predicting the Summer season, 9 of the top 10 features were human pathogens. Among these, Salivirus FHB, Salivirus A and Kobuvirus sp. showed positive associations, whereas Mamastrovirus 1, Norwalk virus, Human astrovirus, Astrovirus MLB1, Sapporo virus showed negative associations with the season. The Tobacco ringspot virus was the most informative feature, with higher values in the Summer (negative correlations). The full list of informative features for binary classification for each individual month and season are shown in Supplementary Table 6 and the top ten informative features are shown in Supplementary Figure 9.

We summarized the union of the top 10 features from regression models trained with one month of history for each of the 14 species (Supplementary Table 8A), and from classification models across all 12 months and 4 seasons (Supplementary Table 8B). Among these top recurrent features for the 14 species, there are 49 human pathogens (78 positive and 22 negative associations) and 14 non-human pathogens (22 positive and 4 negative associations) (Supplementary Figure 9E). Similarly, among the recurrent features from the binary classifiers for each seasonal and monthly predictor (16 models built in total), there are 42 human pathogens (75 positive and 40 negative associations) and 16 non-human pathogens (33 positive and 11 negative associations) (Supplementary Figure 9F).

As outlined above, we built two regression models for month number and season number. The predictor for month number achieved a median R^2^ of 0.75 (Figure 5H), and the predictors for season number achieved an R^2^ of 0.64 (Figure 5I). The full lists of top informative features for month and season number are in Supplementary Tables 7A,B. Among the top 10 important features for the month predictor, 8 were human pathogens; Sapporo virus and Primate bocaparvovirus 1 were positively associated, whereas Human respirovirus 3, Rotavirus A, Mamastrovirus 1, Influenza A virus, and Human astrovirus were negatively correlated with month number. For the season predictor, 8 of the top 10 features were human pathogens—Human astrovirus, Human Respirovirus 3, Respiratory syncytial virus, Rotavirus A, Rhinovirus C, Mamastrovirus 1, and Norwalk virus, all negatively associated with season number. Tobacco ringspot virus and squash mosaic virus were two plant viruses among the top 10 features and showed positive associations with both month and season numbers. The potato, tobacco, and squash mosaic viruses could be related to dietary or recreational habits of the population; since pets are an important part of life for many, viruses such as Feline Rotavirus could show seasonal patterns of disease for pets that might be otherwise underappreciated.

## DISCUSSION

This study presents, to our knowledge, the most comprehensive temporal and spatial analysis of the human wastewater virome to date, looking at three years of continuous metagenomic data spanning hundreds of viral species across 15 Texas cities. Our results demonstrate three key insights: 1) the human virome exhibits robust, predictable seasonality far beyond well-characterized respiratory virus peaks, 2) viral species display complex co-occurrence networks that vary by season and, perhaps most importantly, 3) historical virome data can successfully predict future pathogen abundance with operationally useful accuracy. These discoveries have immediate implications for public health and outbreak preparedness.

Perhaps the most striking finding is the consistent seasonality of the human virome. While seasonal patterns in respiratory viruses like influenza and RSV are well-documented, our data suggests that seasonality is a fundamental organizing principle across the entire detectable virome, encompassing 76% of genera spanning diverse viral families and routes of transmission. The organization into three distinct seasonal clusters, with viruses peaking in Spring/Fall, Summer, or Winter, suggests that common environmental, behavioral, or host factors drive viral dynamics beyond the specific biology of individual viruses. For Winter-peaking viruses, decreased humidity, indoor crowding, and reduced UV exposure are established drivers (Bloom-Feshbach, Alonso et al. 2013, Qi, Liu et al. 2022, Zhou, Yang et al. 2022). In contrast, the strong Summer signal observed for enteric viruses (Enterovirus, Parechovirus, Kobuvirus) may reflect increased recreational water activities and changes in dietary habits (Sinclair, Jones et al. 2009, Ayukekbong, Andersson et al. 2014, Lanrewaju, Enitan-Folami et al. 2022). The intermediate cluster which peaks during Spring and Fall with Summer lulls, may represent viruses sensitive to temperature extremes or those benefiting from patterns in school attendance. Notably, plant and arthropod viruses exhibited defined seasonal peaks, suggesting that environmental virome seasonality driven by agricultural cycles and vector biology is captured alongside human-shed viruses, underscoring wastewater as a unique sentinel for both public health and ecological dynamics. Our work confirms the high abundance of these plant-based viruses in WW (Duarte, de Andrade et al. 2023, Cuevas-Ferrando, Sánchez et al. 2025, Worp, Nieuwenhuijse et al. 2025) and we were able to quantify the seasonal patterns of such viruses.

The fact that these seasonal patterns were observed consistently across cities separated by over 700 miles is remarkable and suggests that drivers of virome seasonality operate largely independent of at least regional geographic scales. This reinforces the possibility that the results reflect synchronous population-level behaviors across Texas or climate patterns that impose similar selective pressures on viral transmission despite local differences in weather. Practically, this consistency suggests that predictive models trained in one location may generalize to others within the same region, potentially allowing resource-limited jurisdictions to pool resources or benefit from monitoring conducted in neighboring areas.

The correlation network analysis showed complex associations between viral species, with the majority representing human-human pathogen pairs. These correlations are likely attributable to multiple mechanisms including co-infections, shared seasonal drivers, or viral reactivation cascades where one infection triggers immunosuppression enabling reactivation of latent viruses. The relative lack of negative correlations is striking and suggests that competitive exclusion between viruses is rare, at least at the population level despite being well-established in experimental systems. There are a few potential explanations for this such as most viruses infecting different cell types, population-level dilution effects obscuring individual-level interference, or our sampling intervals being too coarse to capture transient dynamics. Importantly, additional correlations within seasonal clusters reveal that viral interactions are context-dependent, suggesting predictive models should account for time of year because associations relevant in one season may not hold in another.

Our successful prediction of pathogen abundance one month in advance represents a significant step towards operational disease forecasting. More than 50% of modeled species achieved R^2^>0.50 with modest historical data, while ∼10% reached R^2^>0.75 with minimal training. This is outstanding performance considering the stochasticity in viral transmission and the complexity of factors influencing pathogen shedding. Species with immediate predictability like Rotavirus A and SARS-CoV-2 likely exhibit strong transmission momentum and temporal autocorrelation, meaning once prevalent, they remain prevalent for extended periods. In contrast, species requiring extended historical training data like Influenza A may undergo more volatile cycles driven by antigenic drift or stochastic effects, requiring longer temporal context to capture multi-year patterns. Additionally, the finding that predictive features often include other viral species rather than the target virus itself has profound implications. It suggests the virome functions as a complex interconnected ecological system where one virus’s abundance provides information about the entire community. This network effect strongly suggests that comprehensive virome monitoring provides more predictive power than single-pathogen monitoring.

The performance in predicting sample collection season (AUROC >0.95) demonstrates that the virome operates as a “biological clock” synchronized to environmental and behavioral rhythms. This could be utilized for temporal calibration of samples with unknown collection dates or for quality control in monitoring programs. More importantly, however, the top predictive features could potentially serve as sentinel indicators for seasonal transitions, enabling early warnings and dynamic adjustments of monitoring intensity.

There are, of course, limitations to this study to consider. Our analysis focused on Texas cities with similar, but not identical, climates. Validation in geographically and climatically diverse regions is needed to assess generalizability of these methods. Our sampling frequency may also miss transient viral spikes, and the nature of observational data limits causal inference about mechanisms driving seasonality and co-occurrence correlations. Incorporating external covariates like temperature, humidity mobility data, and vaccination coverage could improve predictions and enable mechanistic interpretation. These improvements are currently being investigated. The detection of non-human viruses also raises interesting questions about their sources. While some likely represent dietary intake or environmental contamination, systematic investigation could yield insights into connections between human, animal, and environmental health. Finally, prospective validation is essential; while our retrospective approach provides evidence of predictive skill, real-time implementation will determine operational utility. We are currently deploying a pilot forecasting system in select Texas cities to assess real-world performance.

This work demonstrates that the human wastewater virome is not a chaotic soup of opportunistically shed pathogens, but rather a structured, seasonal ecosystem. The pervasive seasonality, intricate co-occurrence networks, and surprising predictability from historical data collectively suggest that viral transmission dynamics at the population level are more predictable than previously appreciated. This predictability appears to provide the opportunity to monitor the virome comprehensively and apply machine learning to detect patterns that can be used to forecast pathogen activity before outbreaks occur, potentially helping to shift disease monitoring from reactive to proactive. As wastewater-based epidemiology becomes more established globally, the integration of metagenomic sequencing and predictive analytics positions this approach as a cornerstone of modern biomonitoring. The methods and insights presented here provide a foundation for developing operational forecast systems that protect populations from infectious disease threats.

## METHODS

### Virome data collection and processing

Demultiplexed raw FASTQ-format sequences were processed using BBDuk to quality trim (Q25), and remove Illumina adapters. Trimmed FASTQs were mapped to a combined PhiX (standard Illumina spike in) and human reference genome database (GCF_000001405.39) using BBMap to remove human/PhiX reads. Processed reads were run through the EsViriru pipeline (v0.2.3) with default settings. Specifically, reads were aligned to the entire Virus Pathogen Database (v2.0.2) via minimap, and alignments were filtered by CoverM (Aroney, Newell et al. 2025) (https://github.com/wwood/CoverM) to require at least 90% average identity across 90% of the read length. Virus genomes/segments with reads covering at least 1000 nucleotides or 50% of the genome/segment length were considered preliminary detections. Consensus sequences from this preliminary set of detected virus genomes/segments were extracted using samtools consensus. After removing strings of ambiguous N’s, the preliminary consensus sequences were compared to each other pairwise using anicalc, and clusters were made with sequences of at least 98% average identity using aniclust (Nayfach, Camargo et al. 2021). The longest consensus sequence in each cluster was kept as the final sequence. The sequencing reads were then aligned to a smaller database of only the references corresponding to final sequences with the same parameters for minimap2/CoverM. Virus genomes/segments from this alignment with reads covering at least 1000 nucleotides or 50% of the genome length were considered final detections. The metric RPKMF was calculated as (Reads Per Kilobase of reference genome)/(Million reads passing Filtering).

Virome abundance data were provided as RPKMF at multiple taxonomic levels. For the analysis presented here, the abundance at species and at genera level was utilized. Overall, virome abundance profiles were computed for 2908 samples spanning 36 months, from May 2022 to April 2025.

### Data processing and clustering

Virome abundance data in RKPMF was processed using the Scanpy library (Wolf, Angerer et al. 2018) and converted to log2(RPKMF+1). Uniform Manifold Approximation and Projection (UMAP) representations have been used for microbiome data analysis (Armstrong, Martino et al. 2021). Based on its computational efficiency, principal component analysis, neighborhood detection, and UMAP representations were computed using the Scanpy package (Wolf, Angerer et al. 2018). Sample clustering was implemented using the Leiden community detection algorithm (Traag, Waltman et al. 2019). Batch effect was computed using the k-nearest-neighbor batch-effect test (k-Bet) approach (Büttner, Miao et al. 2019). Sample cluster signatures were determined using a non-parametric Spearman rank test, as implemented in Scanpy. Hierarchical clustering tree across different resolutions was implemented using the clustree R package (Zappia and Oshlack 2018). Species and genera distribution over time were plotted using the GraphPad system v10.6.1 and the Python scientific library.

### Computing species and genera correlations

For each pair of species and each pair of genera, we computed non-parametric Spearman rank correlation. Multiple hypothesis testing corrections were performed using the Benjamini-Hochberg test for false discovery rate (FDR), with significance considered at FDR<0.05 and r^2^>0.2. We considered human and non-human pathogens using multiple references including the Viral Diseases Explorer tool available through the Virus World database (https://doi.org/10.1128/jvi.00620-23) (Rojas Labra, Montiel-Garcia et al. 2023) and NCBI Virus Sequence Similarity Search Interface (VSSI) (National Center for Biotechnology Information (NCBI) 2004) (https://www.ncbi.nlm.nih.gov/labs/virus/). Species and genera distribution over time and scatterplots were plotted using the GraphPad system v10.6.1 and the Python scientific library.

### Predictive models using species relative abundance

We applied machine learning (ML) random forest (RF) to build separately binary predictors (classifiers) and numerical predictors (regressors), using the PyCaret (Moez A. 2020) and sklearn (Pedregosa, Varoquaux et al. 2011) Python implementations. ML models were built over 100 cross-validation iterations as follows: for each predictor, we split the data into 70% training and 30% testing, then performance metrics were computed. For classification, the performance metric used was area under receiver operating characteristic curve (AUROC), whereas for regression the performance metric used was goodness-of-fit R^2^. For each predictor, the distribution of performance metrics over 100 cross-validation iterations was collected, then the final median value was reported. Feature importance was determined using the Random Forest feature importance collected in PyCaret.

### Predictive models for the sample collection month and sample collection season

To build predictors for the month and for the season in which a sample was collected, we set up two types of analysis. 1) **Classification**. We encoded each month using one-hot encoding, as follows: January or not-January (e.g. February to December), February or not-February, and so forth. We encoded each season as Winter or not-Winter (e.g. Spring, Summer, or Fall), Spring or not-Spring, and so forth. This approach set up 16 classification problems. 2) **Regression.** We encoded each month as a number, with January as 1, February as 2, and so on through December as 12. Next, we encoded each season number, with Winter as 1, Spring as 2, Summer as 3, and Fall as 4. This approach set up 2 numerical regression problems. For both classification and regression, we used all samples each month per site (n=754) for training and testing, using the machine learning classification and machine learning regression approaches described above.

### Predictive models for virus levels one month in the future

To predict the level of virus levels for one virus one month into the future, we considered 1 to 12 months of history. For n months of history (n ranging from 1 to 12) the input data for each site was set up as follows: 1) for each of the n previous months, the average levels of each species at that site was considered, encoded as “month_minus_1”, “months_minus_2”, …, “month_minus_n” 2) the month and the season of the observation to be predicted were considered as input variables, encoded as outlined above as both binary variable using one-hot-encoding for each month and season, and also as numerical variables in the range 1-12 for months and 1-4 for seasons. Once the problem was set, the machine learning regression approach was applied as described above.

For each species, we trained and tested models using 1 to 12 months history of data, if data was available, resulting in total of 12 regression models per species. Note that for each species and each number of months history, we built a model only when at least 20 sites/mo had nonzero average abundance in training input. With these constraints, data was available to model only 159 species of the overall 374 detected species. Further, for some species in the list of 159 species, there were not enough nonzero data for each site/mo to model each of the 12 months of history, in general with fewer species having a larger number of history data available.

## Data Availability

All data produced in the present study are available upon reasonable request to the authors.

## ACKNOWLEDGMENTS

This work was in part supported by funding for TexWEB via the Texas Epidemic Public Health Institute (TEPHI), which received funding from the 87^th^ Texas Legislature to operate the Texas Wastewater Environmental Biomonitoring Network, the NIH Genomics Center of Infectious Disease, and seed funds from Baylor College of Medicine. ZV, SLG, TG, and CC were partially supported by the Cancer Prevention Institute of Texas (CPRIT) grants RP170005, RP200504, RP210227, and RP250580, by NIH P30 shared resource grant CA125123, by NIEHS grants P30 ES030285 and P42 ES027725, and by NIH grants S10OD032185, U54CA274321.

## Supplementary Tables Captions

**Supplementary Table 1.** kBet scores showing the average rejection rate in the range of 0 and 1. The smaller value means well-mixed batches.

**Supplementary Table 2.** Specification of 115 genera and 374 species as human or non-human pathogens.

**Supplementary Table 3. A.** Number of samples across each cluster and season. **B.** Significant genera markers for clusters. **C.** Genera markers for clusters before filtering for FDR.

**Supplementary Table 4. A.** Significant species markers for clusters. **B.** Species markers for clusters before filtering for FDR.

**Supplementary Table 5.** ML derived important features for regression modeling of the selected 14 species using one month history.

**Supplementary Table 6.** ML derived important features for seasonal and monthly classification models.

**Supplementary Table 7.** ML derived important features for seasonal and monthly regression models. **A.** Season number as prediction variable by regression model (Winter=1, Spring=2, Summer=3, Fall=4). **B.** Month number as prediction variable by regression model (Jan=1, Feb=2, …, Nov=11, Dec=12).

**Supplementary Table 8. A.** Recurrence of top ten important ML features across 14 virus species. **B.** Recurrence of top ten important ML features across month or season classification.

## Supplementary Figures

**Supplementary Figure 1.**
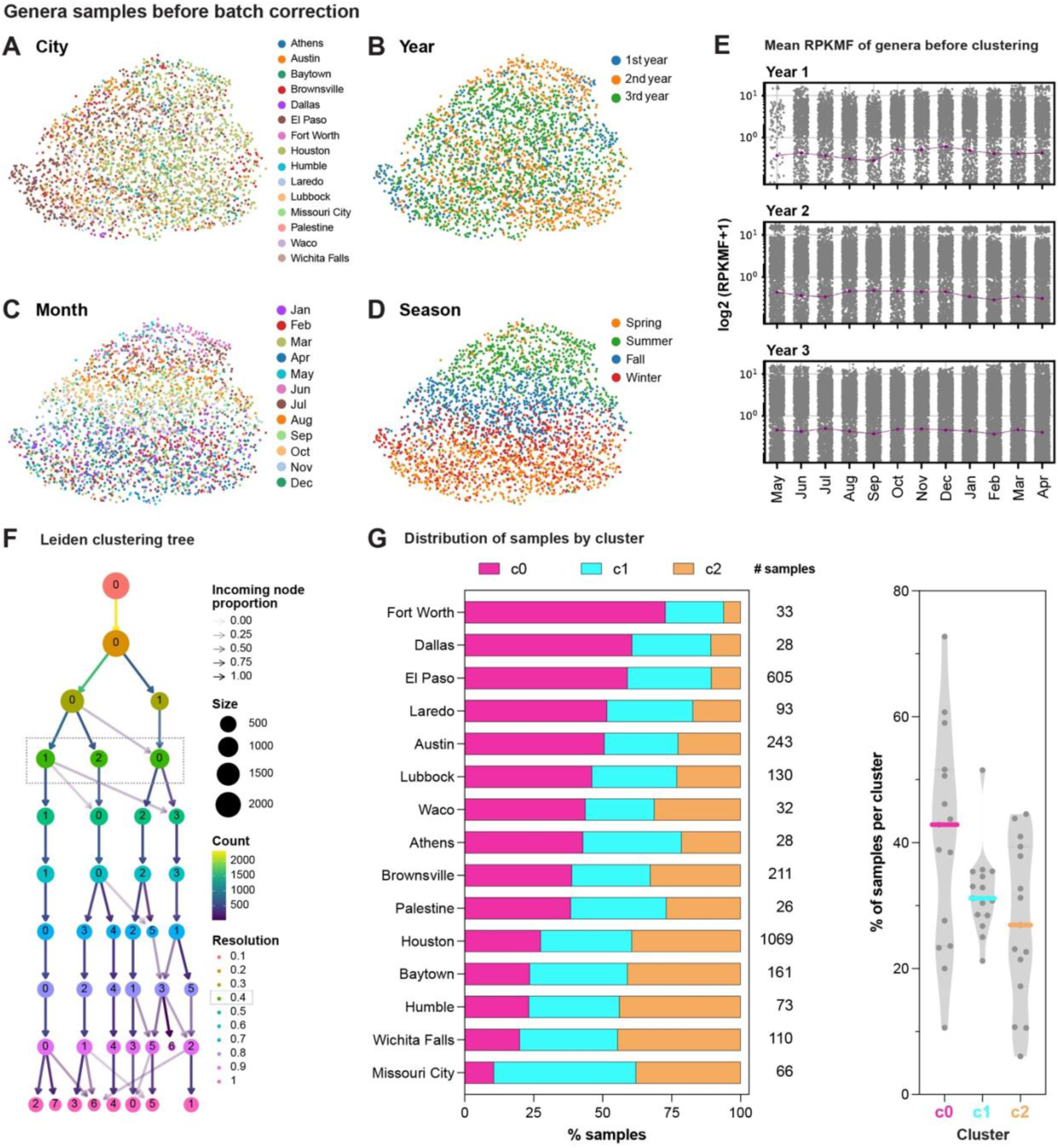
**A-D.** UMAP plots of viral genera showing the organization of samples by: cities, years, months and seasons, before batch correction by city-year. **E.** Mean log-transformed RPKMF of genera across months and years. **F.** Leiden clustering tree showing samples distribution into clusters. **G.** Distribution of samples from 15 cities across three clusters.

**Supplementary Figure 2.**
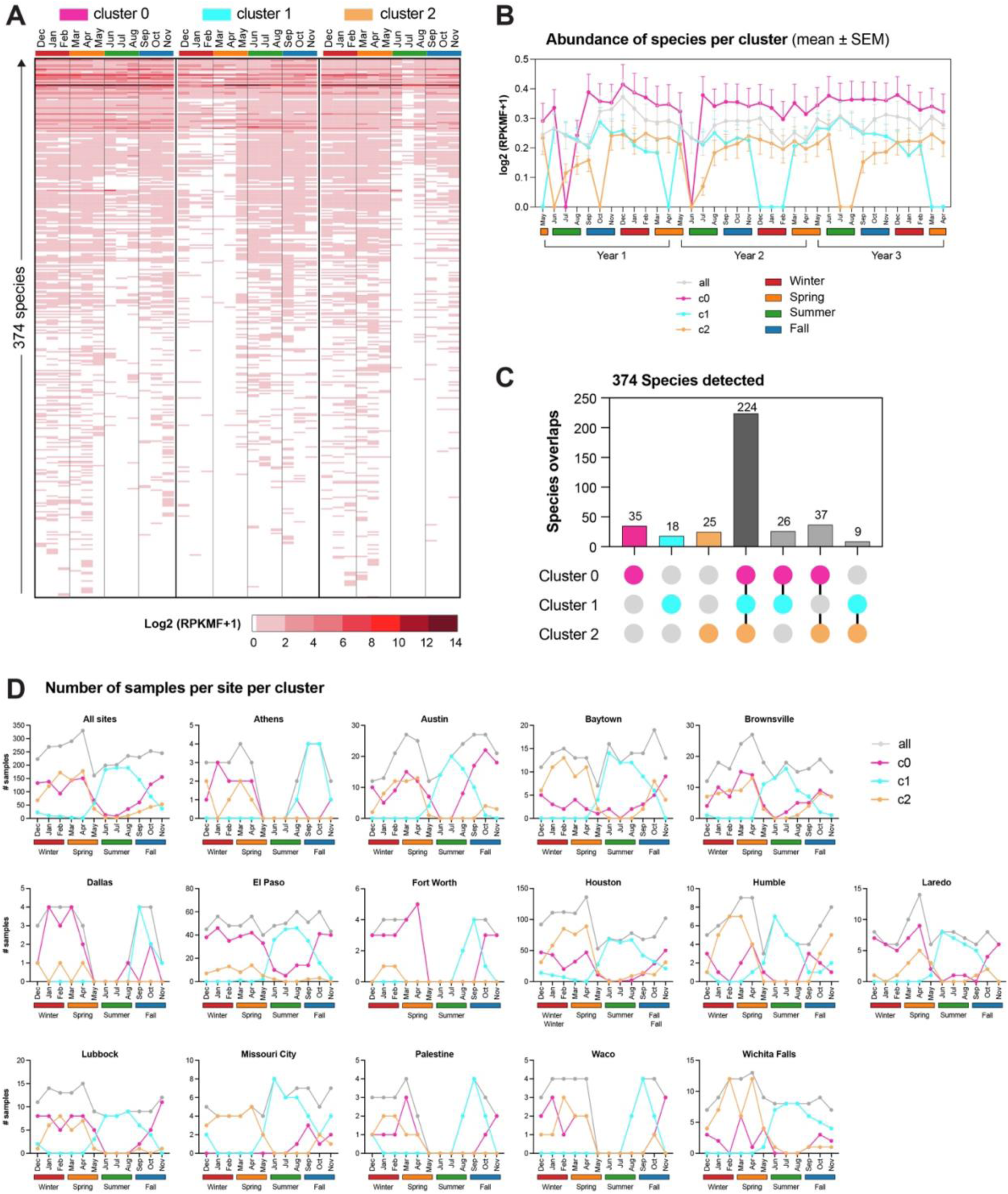
**A.** Mean log-transformed RPKMF values for each of 374 detected species across clusters, seasons and months. **B.** Mean log-transformed RPKMF of species across years, seasons and months. The trend shows monthly averages shown together with standard errors (SEM). **C.** A total of 374 unique species was detected in the samples: 322 species in cluster c0, 277 in cluster c1 and 295 in cluster c2, with 224 shared across all three clusters. The number of species unique to each cluster and those shared between only 2 clusters are shown. **D.** Number of samples per city, grouped by clusters across seasons and months.

**Supplementary Figure 3.**
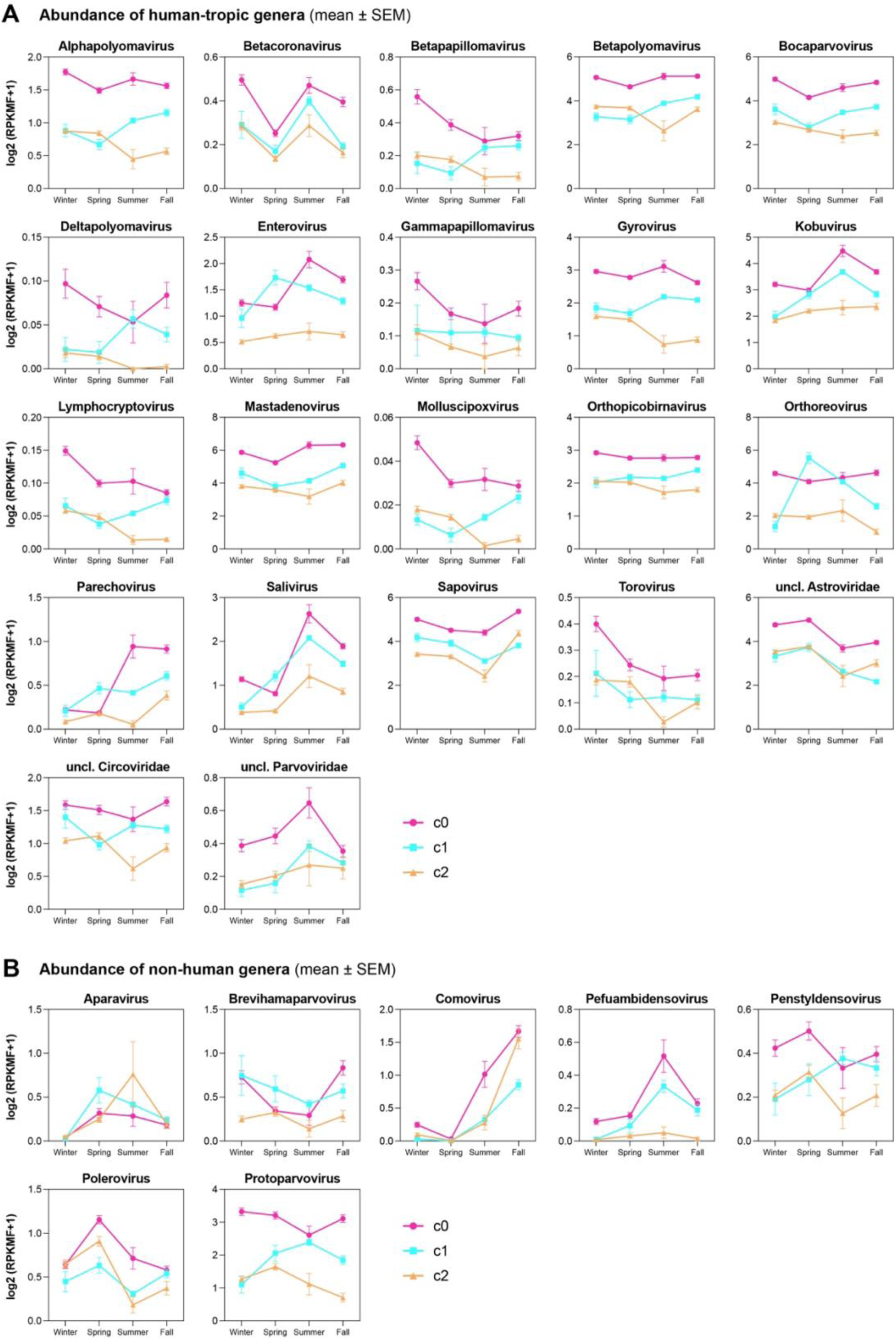
Seasonal distribution of human genera significantly higher in at least two clusters. **A.** Human pathogens. **B.** Non-human pathogens.

**Supplementary Figure 4.**
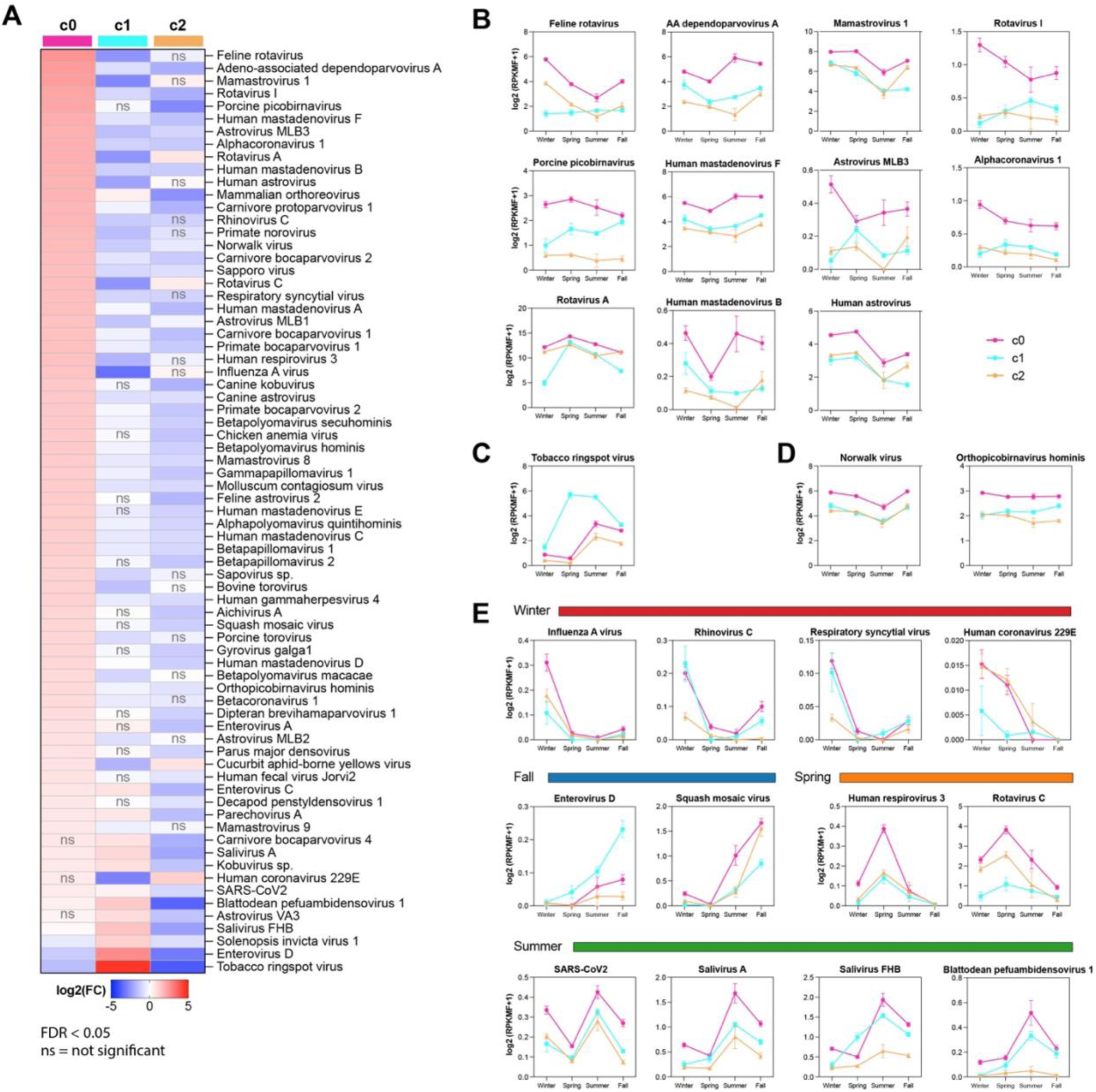
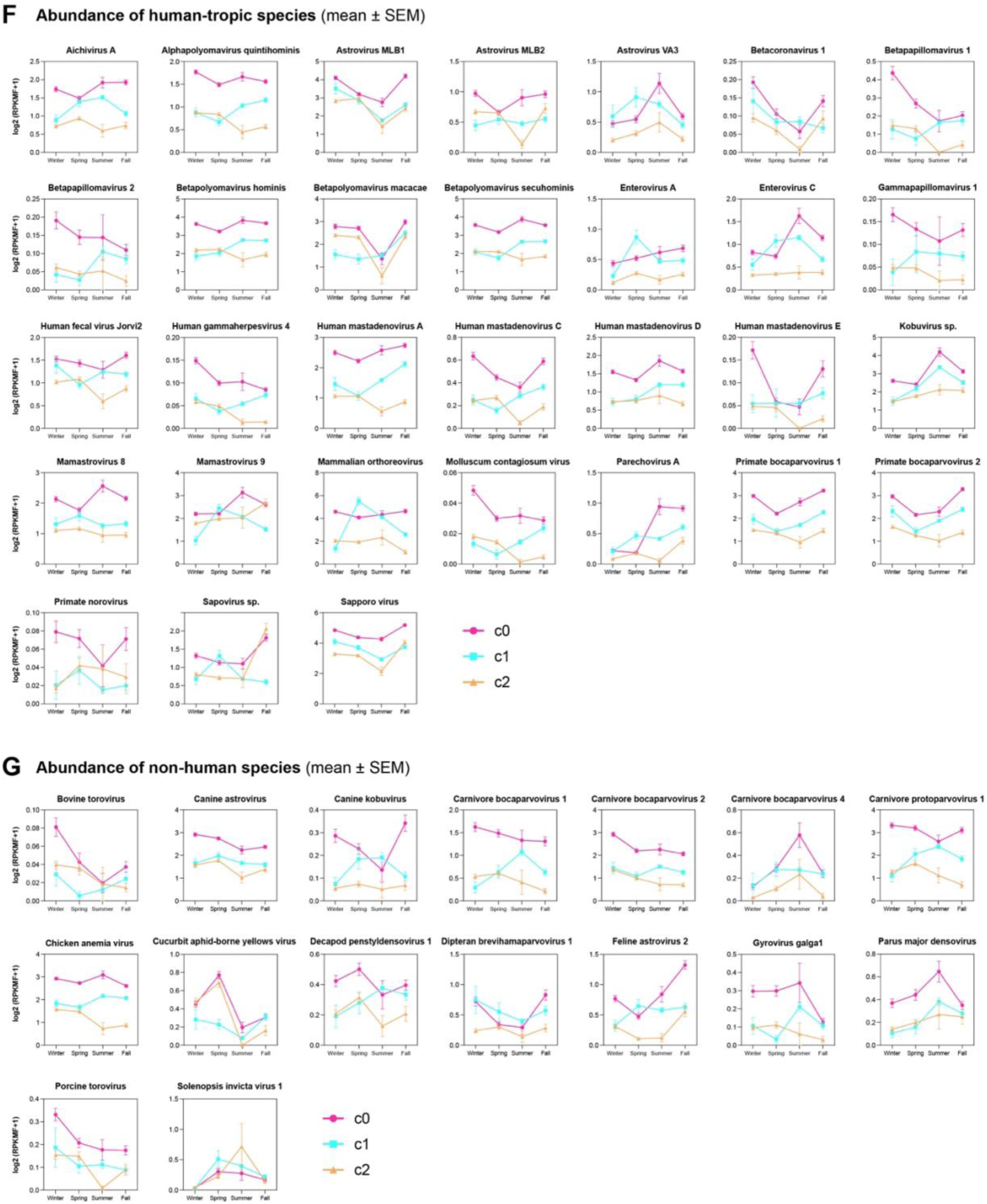
Species markers of the three sample clusters and their seasonal distribution. **A.** Selected significant top species in each cluster (clusters identified at genera level), identified using the Wilcoxon test by comparing log-transformed RPKMF in one cluster against the other two. Significant species selected after filtering for FDR<0.05. **B-D.** Seasonal distribution for selected species. **E.** Seasonal distribution for selected species high in Spring, Summer, Fall, or Winter. **F.** Seasonal distribution of the remaining human species high in any of the three clusters. **G.** Seasonal distribution of the remaining non-human species high in any of the three clusters.

**Supplementary Figure 5.**
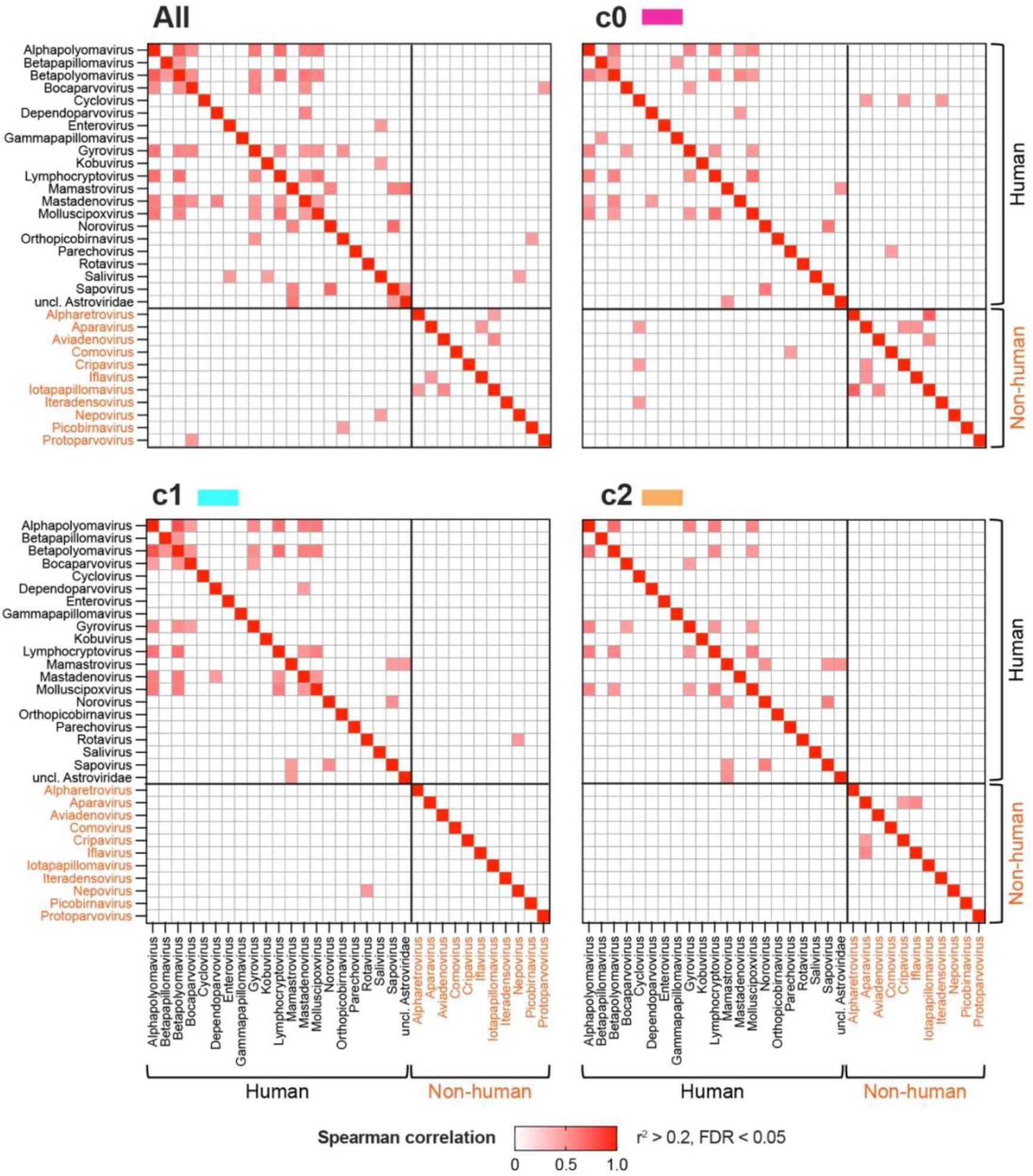
Correlated pathogen pairs at genus level, using all samples, and individual clusters c0, c1, and c2. Data is visualized as correlation heatmaps, split into human pathogens with black font labels, and non-human pathogens with orange labels.

**Supplementary Figure 6.**
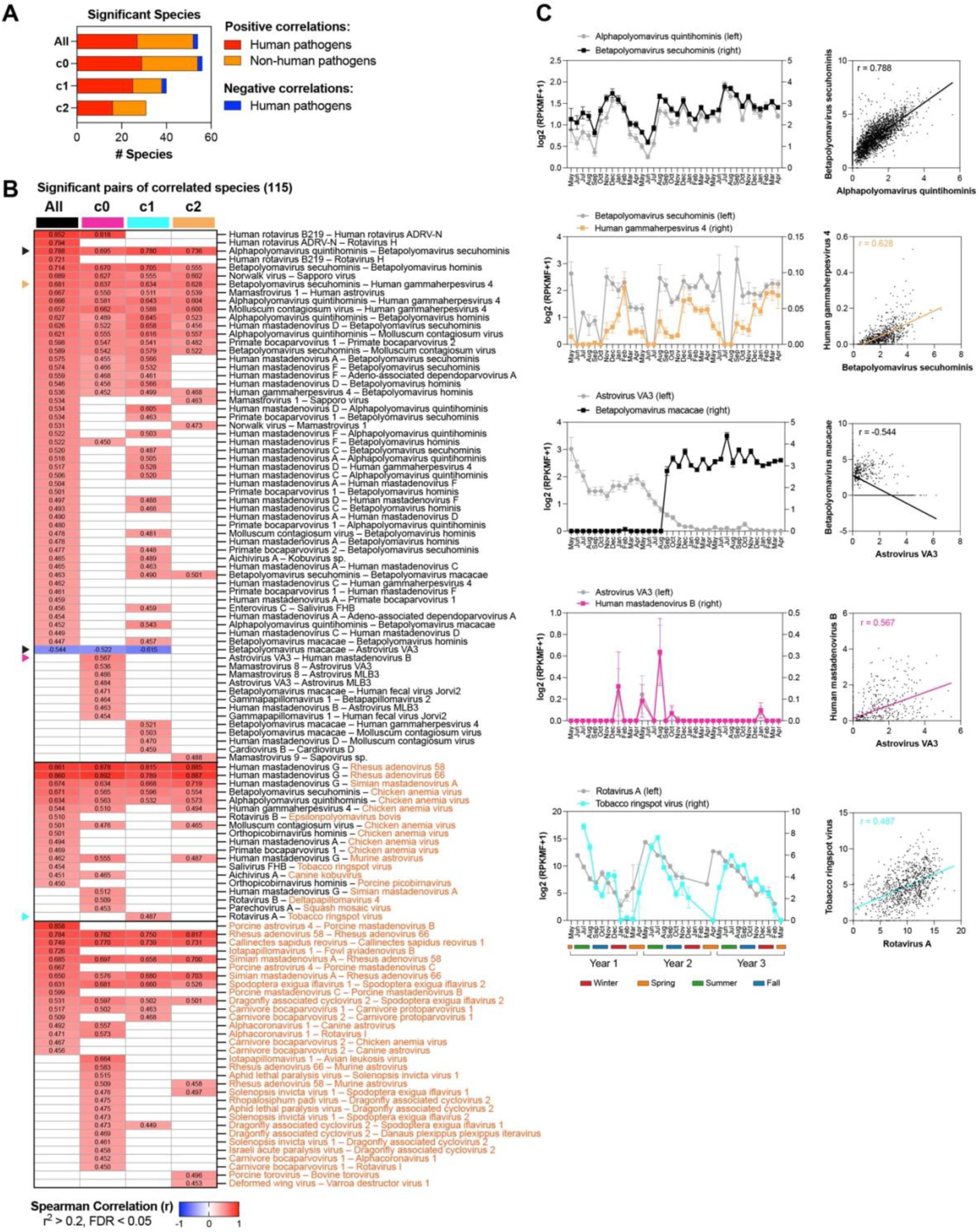
Virome correlation networks at species level resolution. Spearman rank correlation was used to determine correlated species, at FDR<0.05 and r^2^>0.2. **A.** Number of human and non-human pathogen species that show significant correlations using the pooled samples or within each cluster. **B.** Heatmap showing the Spearman rank correlations for significant pathogen species pairs across all pooled samples and in each individual cluster. Human pathogens are indicated with black font, non-human pathogens with orange font. **C.** Visualization of selected correlated pathogen pairs across 36 months and as scatterplots.

**Supplementary Figure 7.**
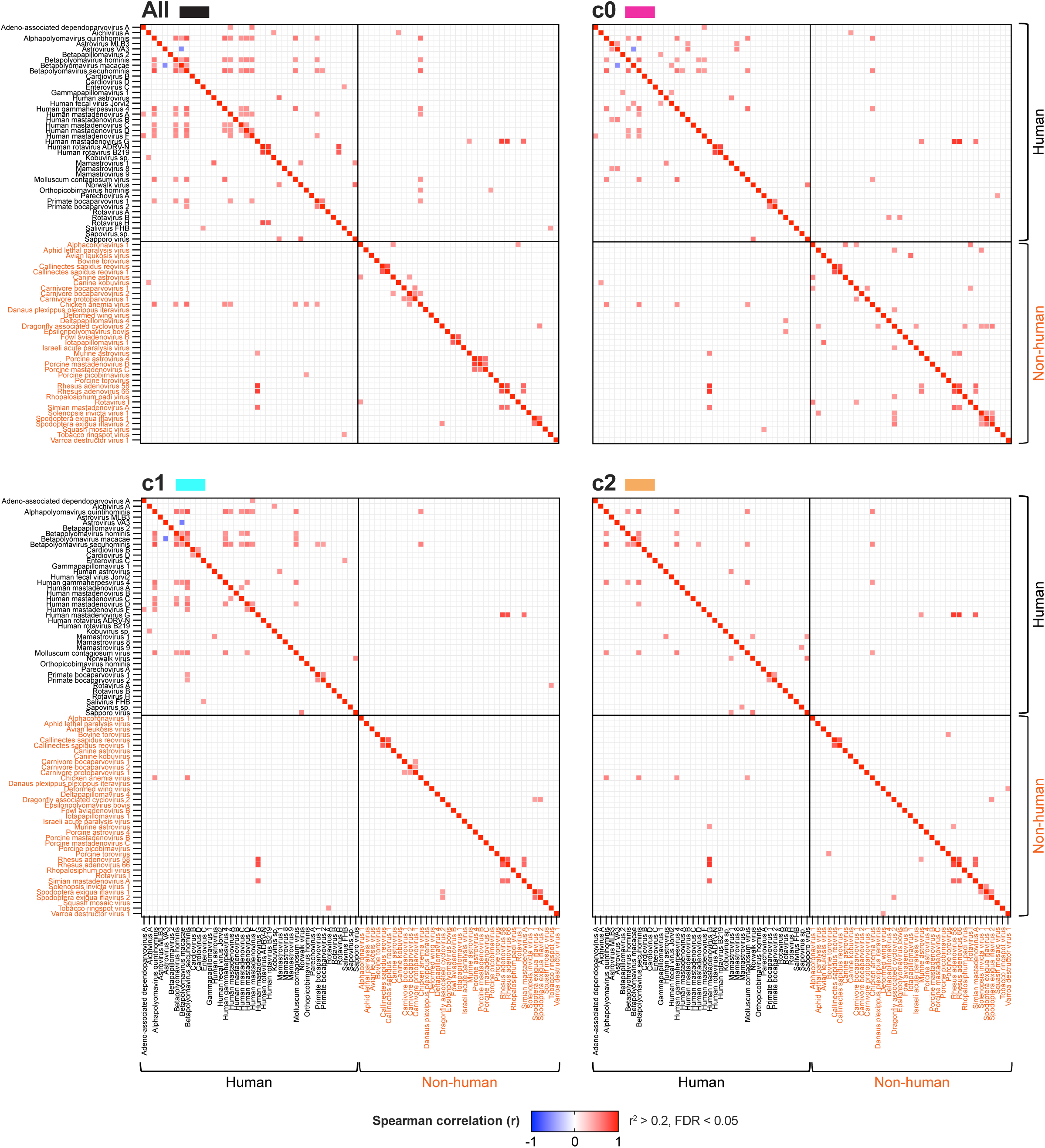
Correlated pathogen pairs at species level, using all samples, and individual clusters c0, c1, and c2. Data is visualized as correlation heatmaps, split into human pathogens with black font labels, and non-human pathogens with orange labels.

**Supplementary Figure 8.**
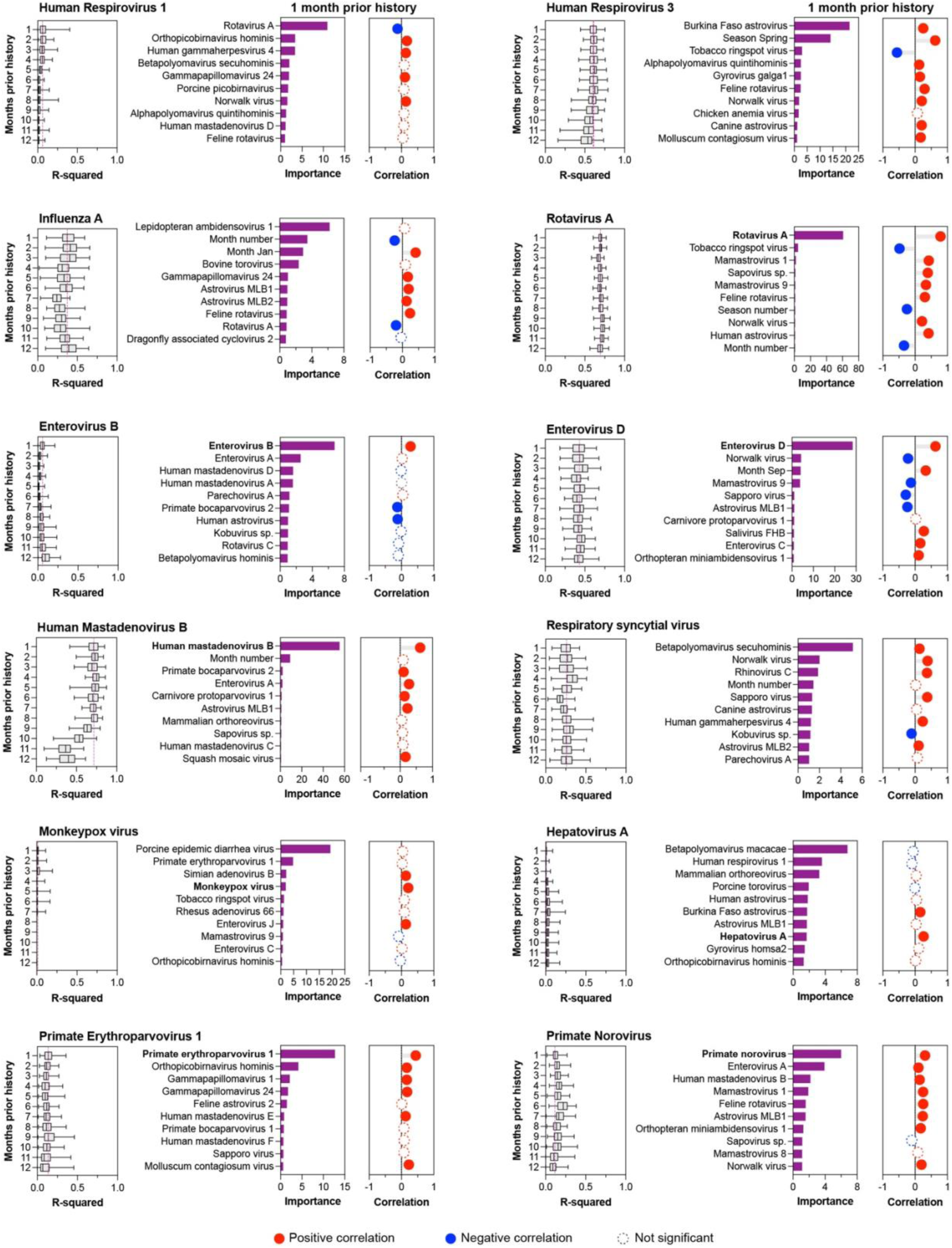
Predictive performance for virus level for 12 species of public interest. For each species we indicate the distribution of R^2^ performance across the 100 cross-validation iterations, the top ten informative features, and direction of association with virus levels.

**Supplementary Figure 9.**
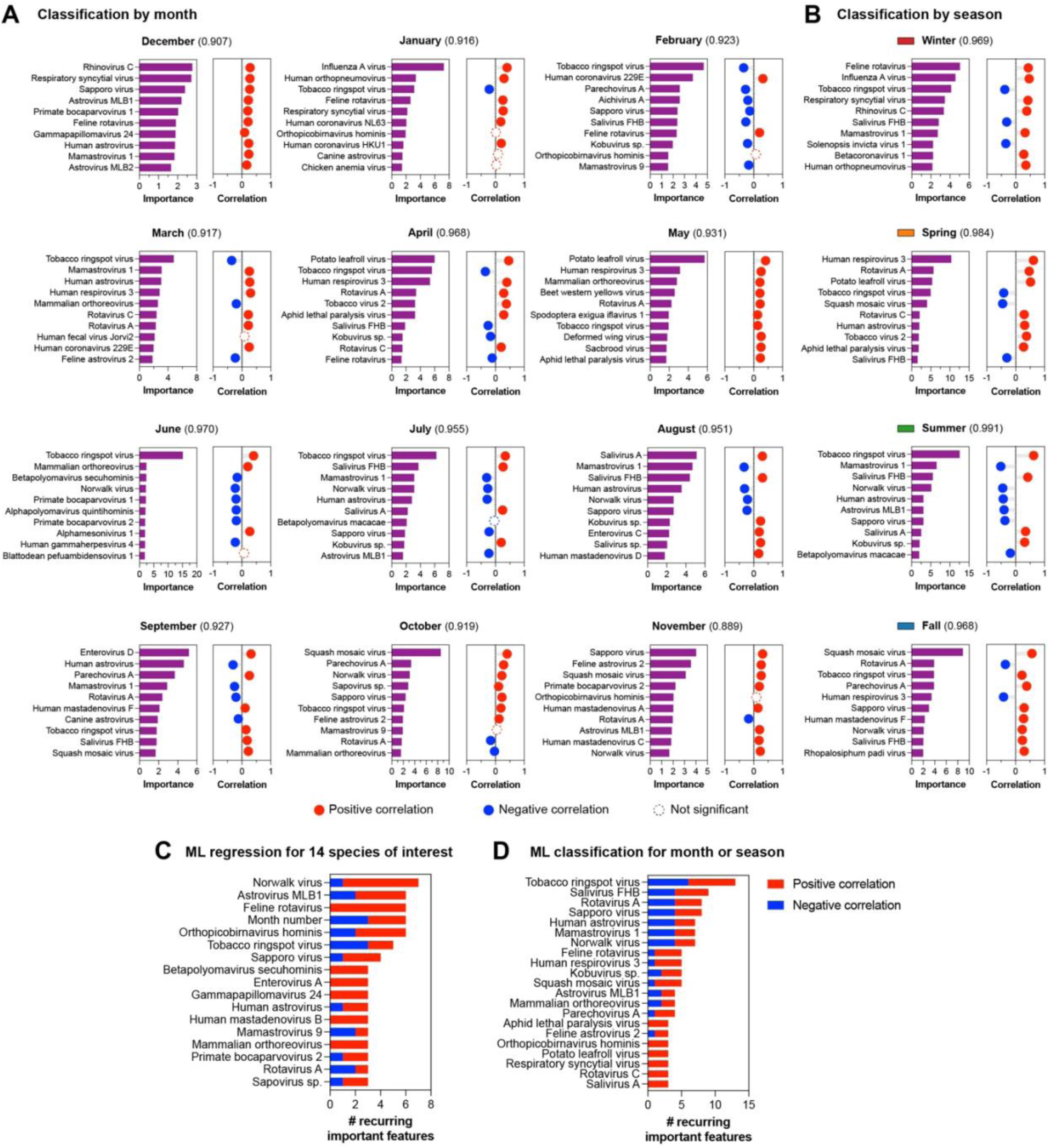
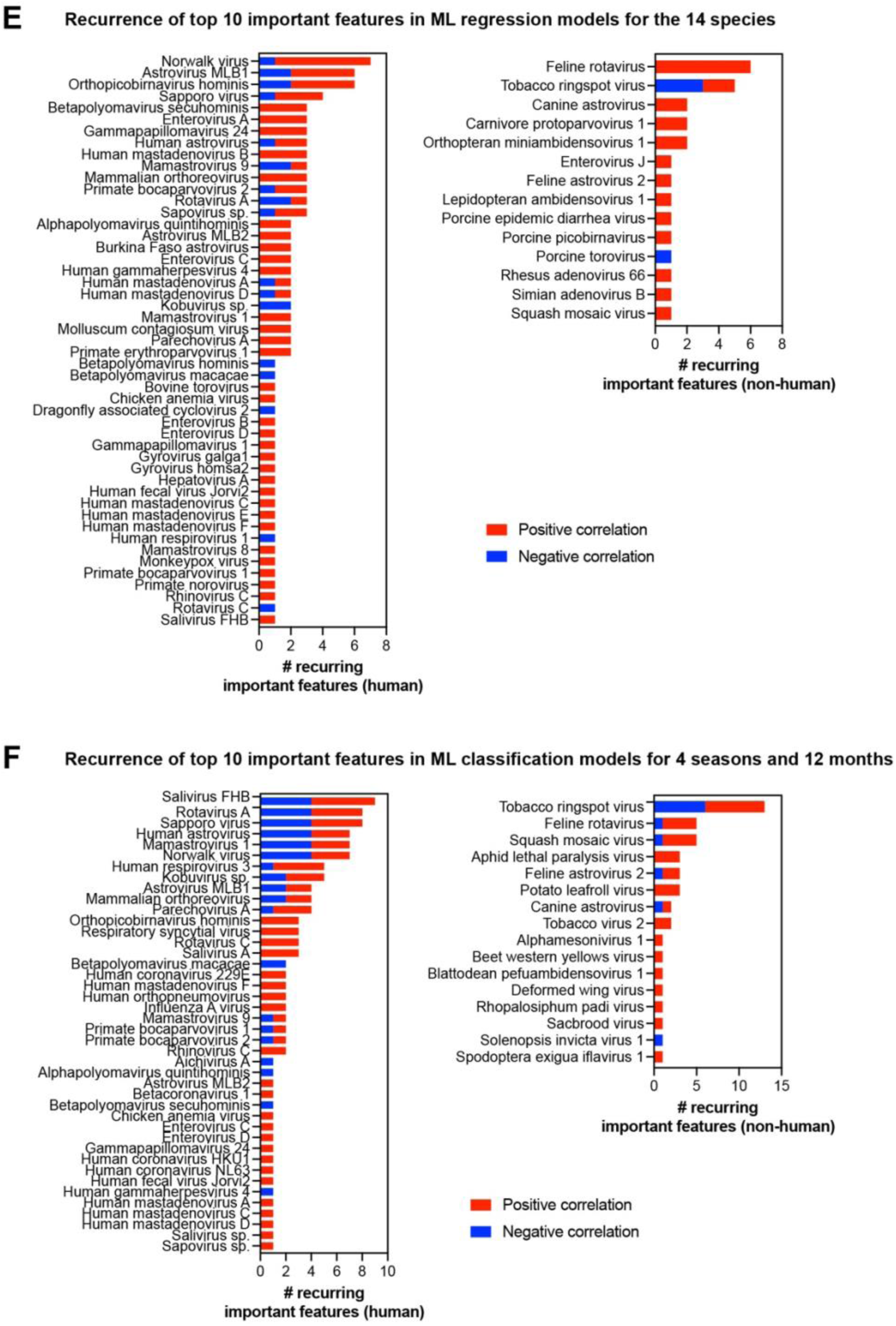
**A-B.** Top ten informative features for individual month and season classifiers, including direction of association. The month of April and Summer season were also shown in Figure 5; added here for completeness**. C.** Summary of top ten informative features recurrent across multiple predictors for the 14 species. **D.** Summary of top ten informative features recurrent across multiple predictors for the months and seasons. **E.** Complete list of the top ten informative features recurrent across predictors, split into human and non-human pathogens for the 14 species. **F.** Complete list of the top ten informative features recurrent across predictors, split into human and non-human pathogens for the month and season classifiers.

